# Childhood cancer in singletons conceived via medically assisted reproduction in Australia: a population-based cohort study

**DOI:** 10.64898/2026.04.08.26350447

**Authors:** A.R. Walker, C.M. Vajdic, A.C. Anazodo, N.F. Hacker, S. Opdahl, M. Chapman, U.M. Sansom-Daly, L. Jorm, R.J. Norman, C. Stern, G.M. Chambers, C. Venetis

## Abstract

1.

**Study question:** Do singletons conceived by medically assisted reproduction (MAR) experience an elevated incidence of childhood cancers and are they at a greater risk of such cancers compared to naturally-conceived singletons?

**Summary answer:** We found no strong evidence the adjusted risk of childhood cancers is increased for MAR-conceived singletons.

**What is known already:** There is longstanding concern children conceived via MAR may be at increased risk of childhood cancer. Current epidemiological evidence does not support such a relationship.

**Study design, size, duration:** We conducted a retrospective population-based cohort study of 5,104,121 singletons born in Australia between 1991 and 2019. Median follow-up time varied from 4 to 10 years depending on mode of conception.

**Participants/materials, setting, methods:** We linked birth records to public medical insurance data of the mother to ascertain MAR conception. We classified treatment as ovulation induction/intrauterine insemination (OI/IUI) or assisted reproductive technology (ART; IVF/ICSI), with ART coded as either fresh embryo transfer or frozen embryo transfer. The cohort included 4,924,354 naturally-conceived singletons and 179,767 singletons conceived via MAR. We calculated standardised incidence ratios (SIRs) to ascertain differences in population incidence of childhood cancer, and generated hazard ratios (HRs) using flexible parametric survival models controlling for key confounders. We report absolute incidence and risk differences for both statistical approaches.

**Main results and the role of chance:** There was no increase in the incidence or risk of all childhood cancers combined for singletons conceived via MAR, either any MAR or specific MAR types. There was some evidence the incidence of leukemias, myeloproliferative diseases, and myelodysplastic diseases was increased after ART compared to the general population (SIR: 1.32, 95% CI 1.02-1.68; equating to 2.09, 95% CI 0.13-4.44 extra cancers per 100,000 person-years), but no increased risk after adjusting for available confounders (HR: 1.04, 95% CI 0.73-1.46). These cancers showed increased incidence and risk for those conceived via IVF (SIR: 1.54, 95% CI 1.01-2.26; HR: 1.77, 95% CI 1.06-2.95), but not ICSI (SIR: 1.27, 95% CI 0.83-1.85; HR: 0.76, 95% CI 0.48-1.22). Incidence of renal tumours was elevated after IVF (SIR: 2.37, 95% CI 1.02-4.67; equating to 1.83, 95% CI 0.03-3.99 extra cancers per 100,000 person-years) and frozen transfer ART (SIR: 2.52, 95% CI 1.09-4.97; equating to 2.12, 95%CI 0.12-5.53 extra cancers per 100,000 person-years), however risk was not elevated after adjusting for available confounders (HR: 1.06, 95% CI 0.47-2.38; and HR: 1.63, 95% CI 0.73-3.61 respectively).

**Limitations, reasons for caution:** We did not have information on parental cause of infertility, which could be a confounder for childhood cancer, although we did adjust for parental history of cancer. For many specific cancer types, fewer than 50 cases were observed in total. Given the number of comparisons reported and closeness of the lower-bound confidence interval to 1, we cannot exclude that a significant association between conception via IVF and leukemias, myeloproliferative diseases, and myelodysplastic diseases reflects a type I error.

**Wider implications of the findings:** Our findings align generally with published meta-analyses on the risk of childhood cancers following MAR conception and reinforce the need for very large studies to increase confidence. Parents who have conceived via MAR and their offspring can be reassured there is not strong evidence the treatments increase the overall incidence or risk of childhood cancer.

**Study funding/competing interest(s):** This work was funded by the National Health and Medical Research Council (NHMRC: APP1164852). Dr ARW declares that their involvement in this work was supported by employment at UNSW Sydney. Prof CMV declares payment to their institution from the National Health and Medical Research Council (APP1164852). Prof NH declares payment to their institution from the National Health and Medical Research Council (APP1164852); royalties and licenses for Berek and Hacket’s Gynecologic Oncology (Walters Kluwer); royalties and licenses for Hacker and Moore’s Essentials of Obstetrics and Gynecology (Elsevier); consulting fees from Darwin Hospital and Gold Coast University Hospital; support for attending the British Gynaecological Cancer Society meeting in Aberdeen, UK, Jun 2023; support for attending the Symposium on Gynaecological Cancer in Budapest, Hungary, Nov 2023; support for attending the International conference of the Rajiv Gandhi Cancer Centre in Delhi, India, Mar 2025; and membership of the Medical Advisory Committee for TruScreen (Australia and New Zealand). A/Prof SO declares that they received payment to their institution from the National Health and Medical Research Council (APP1164852); they received a grant from the European Society for Human Reproduction and Embryology (Open call 2022) including payment to their institution; and that they are a member of the Advisory Board of the Cervical Screening Program in Norway through The Norwegian Institute of Public Health (NIPH), for which they were reimbursed travel expenses to their institution. Prof MC declares support for Theramex European Society for Human Reproduction and Embryology registration and Fertility Society of Australia and New Zealand registration and accommodation. A/Prof USD declares that her involvement in this work was supported via an Early Career Fellowship from the Cancer Institute NSW (ID: 2020/ECF1163) and employment at UNSW Sydney. A/Prof USD also declares payment to their institution from the National Health and Medical Research Council (APP2035240) and the Medical Research Future Fund (APP2032214; APP2038377), and the Australian Research Council (DP240100072) as well as current grants from NSW Health, Prince of Wales Hospital Foundation, and unpaid involvement as an Associate Editor for the “Journal of Psycho-Oncology Research and Practice”. Prof LJ declares payment to their institution from the National Health and Medical Research Council (APP1164852). Prof RJN declares they are the Chair of the Clinical Advisory Committee, Westmead Fertility; External mentor at VinMec hospital; Editorial Editor at the journal “Fertility and Sterility”; and has received funding from the National Health and Medical Research Council (NHMRC) for the NHMRC Centre for Research Excellence in Women’s Health in Reproductive Life (CRE WHiRL).

A/Prof CS declares stock or stock options associated with CSL Ltd, Sigma Healthcare Ltd, Resmed Inc, Medical Developments International Ltd, Vitrafy Life Sciences Ltd, Intuitive Surgical, and Steris PLC. Prof GMC declares payment to their institution from the National Health and Medical Research Council (APP1164852). Prof CV declares payment to their institution from the National Health and Medical Research Council (APP1164852); research grants receive from Merck KGaA and Ferring; payments for honoraria from Merk Ltd, Merk Sharpe & Dohme, Ferring, Organon, Gedeon-Richter for being an invited lecturer in scientific meetings/conferences on multiple occasions as well as member of advisory boards for these companies who have a commercial portfolio in the field of assisted reproduction technology (ART); and speaking fees from IBSA, Vianex, Sonapharm; travel support for their participation in scientific meetings/conferences both nationally and internationally, usually as an invited speaker for the following companies – Merck Ltd, Merck Sharpe & Dohme, Ferring, Organon, Gedeon-Richter; unpaid involvement as a Board member of the Hellenic Society of Fertility and Sterility, Member of the Editorial Board of the journal “Human Reproduction”, Senior Deputy of the Coordination Committee of the Special Interest Group “Reproductive Endocrinology” of the European Society for Human Reproduction and Embryology, Member of the Editorial Board of the journal “F&S Reviews”, Member of the Editorial Board of the journal “RBM Online”, Member of the Editorial Board of the journal “Reproductive Biology & Endocrinology”, Member of the Editorial Board of the journal “Frontiers in Endocrinology”, and Member of the Editorial Board of the journal “Reproductive Sciences”.

**Subject:** Reproductive epidemiology

## 2. Introduction

Medically assisted reproduction (MAR) treatment is increasingly common with up to 10% of children born from these technologies in some countries (Adamson et al., 2025). Though these treatments are generally considered safe, the widespread requires rigorous interrogation of the data to ascertain if there are potential health risks posed to children conceived via MAR. In particular, there is long-standing interest in whether MAR may be associated with increased risk of childhood cancer (Bergh et al., 1999). This stems from concerns about the potential epigenetic effects of treatment (particularly on imprinting) (James and Jenkins, 2018), the elevated levels of oestrogen and progesterone embryos are exposed to during treatment (Kolibianakis et al., 2009, Venetis et al., 2013), and potential effects of the culture medium on embryo development (Chronopoulou and Harper, 2015). These concerns are most pertinent for children conceived via ART, including IVF and ICSI, though risks after non-ART MAR treatments involving fertility medications and IUI have also been explored (Brinton et al., 2004, Hargreave et al., 2019).

Recent large-scale and high-quality epidemiological studies of associations between MAR and childhood cancer have generally suggested no relationship between the two, with some variability depending on treatment and cancer type. A Norwegian study of 2.25 million children found no increase in the overall number of childhood cancers for either fresh or frozen embryo transfer (Oakley et al., 2025), replicating findings from a French study of 8.53 million children (Rios et al., 2024). However, a recent Taiwanese study of 2.3 million children found an increased risk of childhood cancers (hazard ratio, 1.58; 95% confidence interval: 1.17-2.12) for children conceived via ART compared to naturally-conceived children (Weng et al., 2022). Similarly, studies from the United States (hazard ratio: 1.31; 95% confidence interval: 1.08-1.59) and the Nordic countries (hazard ratio: 1.65; 95% confidence interval: 1.15-2.20) have shown evidence of increased cancer risk for children conceived via ART, though no consistency with respect to embryo transfer type (Luke et al., 2022, Sargisian et al., 2022). One Dutch study of 82000 children also found an increased risk of cancer in those conceived via ICSI (but not ART generally) when compared to those conceived to women with fertility issues but without the use of ART (hazard ratio: 1.58; 95% confidence interval: 1.08-2.31) (Spaan et al., 2023). Adding to the complexity, some population-based studies have shown increased incidence or risk of specific types of childhood cancer, including leukemias, myeloproliferative diseases, and myelodysplastic diseases; bone tumours; hepatic tumours; embryonal tumours; and central nervous system tumours (Hargreave, et al., 2019, Luke, et al., 2022, Rios, et al., 2024, Sargisian, et al., 2022, Spector et al., 2019, Weng, et al., 2022, Williams et al., 2018).

Despite growing international evidence, there have been no Australian population-level studies examining the risk of cancer in children conceived via MAR and childhood cancer. Australia has one of the world’s highest IVF use per-capita (Peipert et al., 2023), and boasts strong linked-data infrastructure enabling accurate identification of both MAR conception (Choi et al., 2022) and cancer (Australian Institute of Health and Welfare, 2024) over an extended period. As such, Australia provides an ideal setting to examine this question. Our study aimed to describe the population incidence and estimate the association between MAR conception (both overall and by type) and childhood cancer using a large retrospective Australian population-based cohort of over five million singleton children born between 1991 and 2019.

## 3. Methods

### Ethical approval

The study was approved by all applicable human research ethics committees (HRECs) including the Australian Institute of Health and Welfare (AIHW) HREC (EO2019/5/1061). Data access was approved under a waiver of informed consent and researchers accessed linked anonymised data.

### Study design

This study was a retrospective population-based cohort study.

### Cohort definition and formation

#### Data sources and linkage

We derived the primary cohort using record linkage performed by the AIHW in conjunction with state-based Data Linkage Units as described in Vajdic et al. (2026). A brief description of all datasets and their usage can be seen in Supplementary Table 1. Singleton children born between 1 January 1991 and 31 December 2019 to women aged 18-55 years were eligible and identified from Australian jurisdictional perinatal data collections and Registries of Birth Deaths and Marriages. The perinatal data collections included pregnancies reaching either 400 grams birthweight or 20 weeks gestation, with some minor variations between jurisdictions. Children were linked to their registered parents using Registry of Births, Deaths and Marriages datasets. These triads were linked to the Medicare Enrolment File to identify: parents who had not used Medicare services, those with parents who immigrated to Australia, and key demographic variables at the time of birth of the child.

Medicare is Australia’s universal public health care insurance scheme for citizens and permanent residents, with registration occurring soon after birth or residency. This file was linked to the Medicare Benefits Schedule and Pharmaceutical Benefits Scheme datasets to identify mothers who had a claim for a reimbursed MAR-related healthcare service or MAR-related medication from 1 January 1991 to 31 December 2019. As pharmaceutical claims data was only available from 1 July 2002, our capture period for clomiphene citrate dispensations began on this date. The study period varied according to the dates of coverage of the perinatal data collections in the Australian jurisdictions. For example, children born in South Australia and Western Australia could enter the study from 1 January 1991 to 31 December 2019, whereas children born in the Australian Capital Territory could enter the cohort from 1 January 1997 to 1 December 2017.

After data cleaning to identify and exclude duplicate and illogical records, we applied exclusion criteria to form the primary cohort (Figure 1). These steps included the exclusion of multiple births (i.e., those with parity greater than 1), which accounted for fewer than 5% of all births. We made this exclusion as, for a substantial number of pregnancies, we could not differentiate the children born in the same birth episode and therefore could not link individual children to cancer outcomes. From this primary cohort, we formed two sub-cohorts for analysis. The first sub-cohort (the standardised incidence ratios cohort, hereafter “SIR sub-cohort”) was defined for the calculation of SIRs. The second cohort (the “Survival sub-cohort”) was defined for survival modelling. The sub-cohorts differed slightly on exclusion criteria. The SIR sub-cohort excluded those whose sex was indeterminate, as sex was required for standardisation. By contrast, we excluded those from the survival sub-cohort who had illogical or missing data recorded in the perinatal data collections (gestational age missing, over 45 weeks; or under 20 weeks; weight under 400 grams or over 7000 grams; or no information on number of prior pregnancies). This approach was taken because these data collections were used to define key covariates for survival modelling.

**Figure 1:**
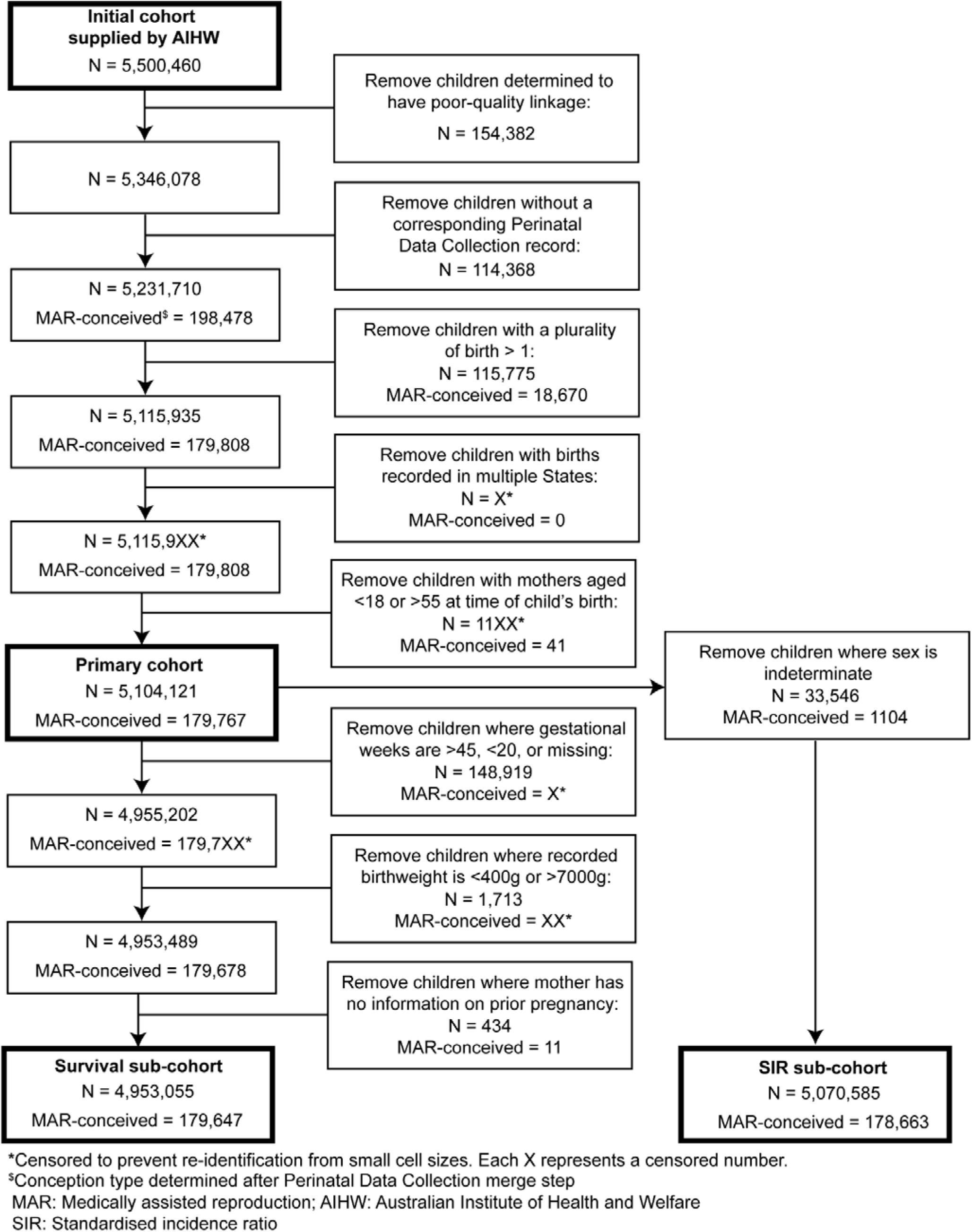
Cohort derivation flow-chart. We first defined the primary cohort, from which we derived two sub-cohorts for analysis

#### Exposure definition

As the vast majority of MAR treatments in Australia are partially publicly subsidised, they are well captured in Australia’s public health insurance scheme datasets, and these data can be used to reliably identify MAR-conceived children in Australia (Choi, et al., 2022). We determined if a child had been conceived via MAR using their mother’s Medicare Benefits Schedule and Pharmaceutical Benefits Scheme data around the estimated time of the child’s conception. We calculated an estimated conception date using estimates of gestational age at birth from the perinatal data collections. We then determined whether the mother had received a MAR treatment during a treatment window either side of that date. We split MAR treatments into OI/IUI or ART treatments. From 2007 onwards, we were able to distinguish the type of ART treatment (IVF or ICSI). ART treatments were coded as either fresh or frozen transfers. Details on calculation of conception date, treatment types and codes, and treatment windows are given in Table 1. We considered all children not conceived via MAR as naturally-conceived. Children conceived via donor gametes were included if their mother received public reimbursement from the Medicare Benefits Schedule for the fertility treatment.

**Table 1:**
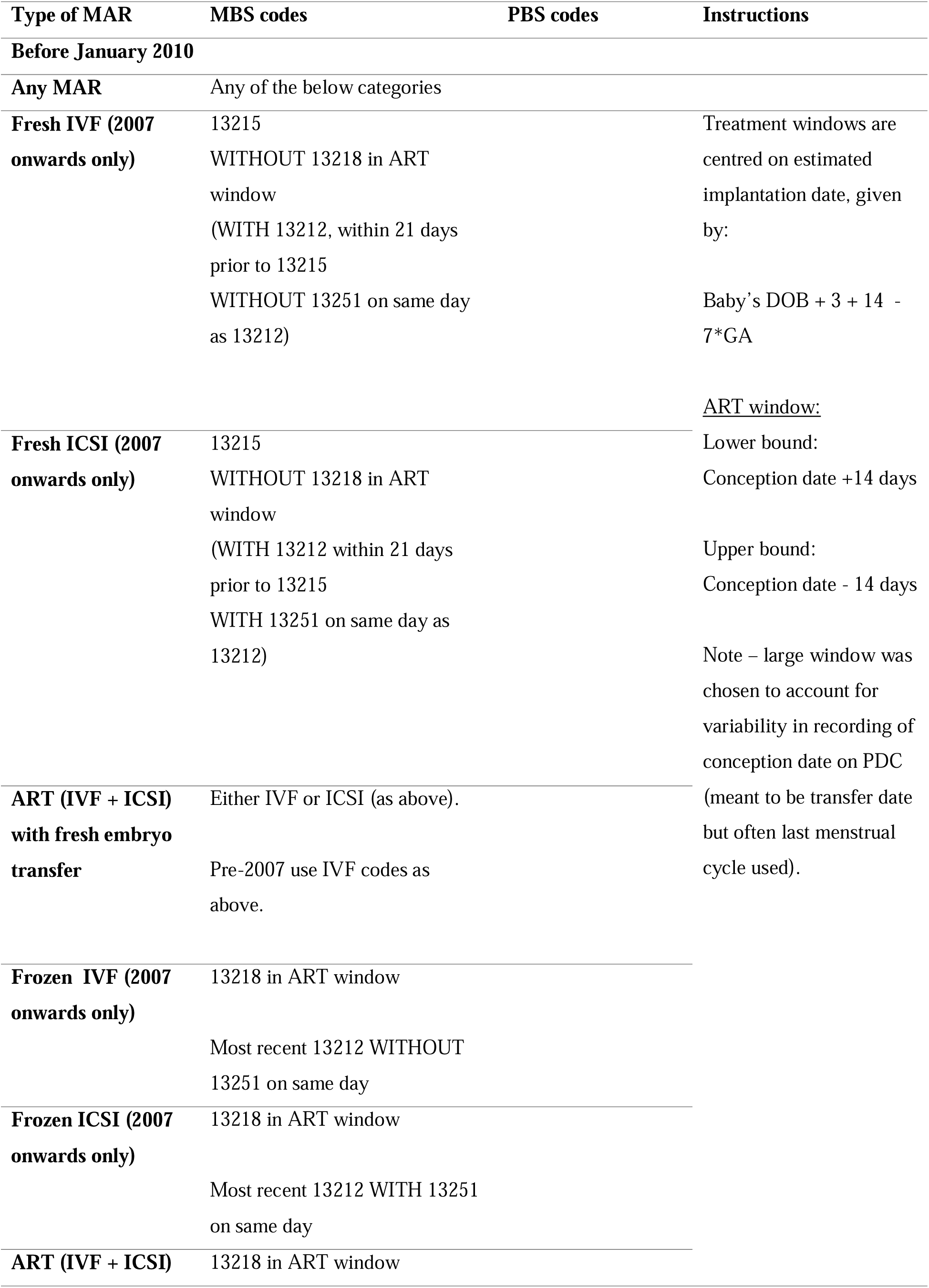
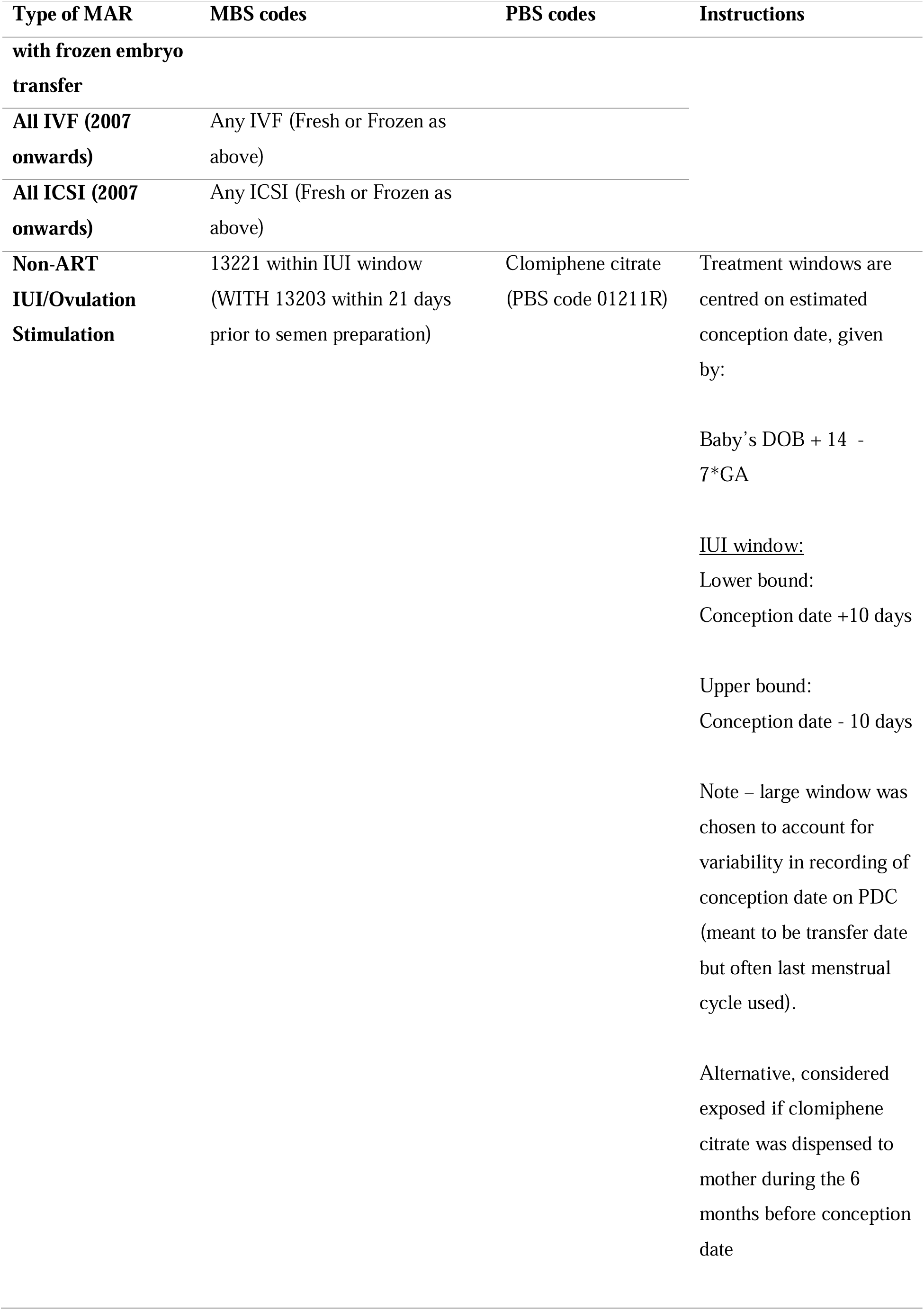
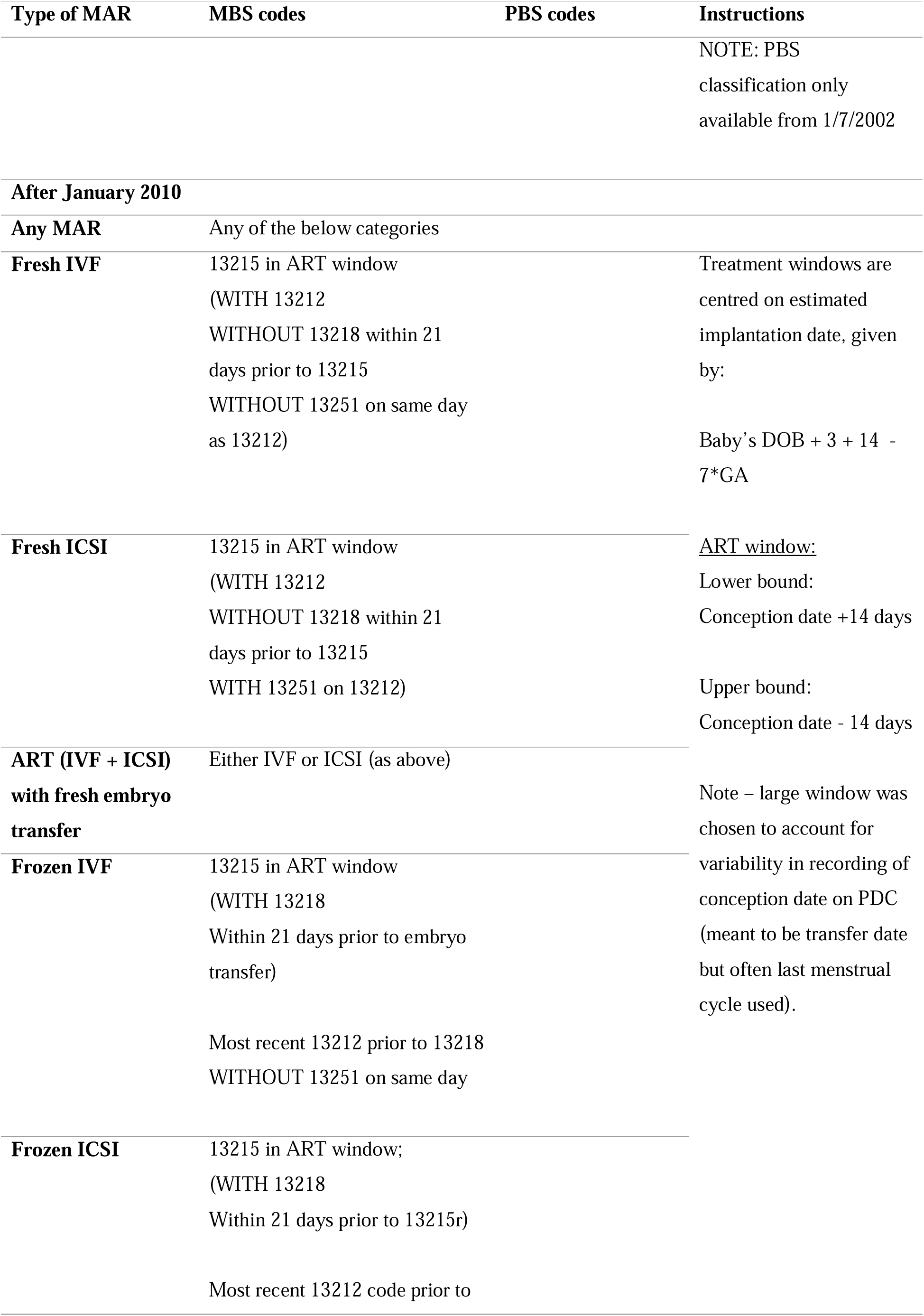
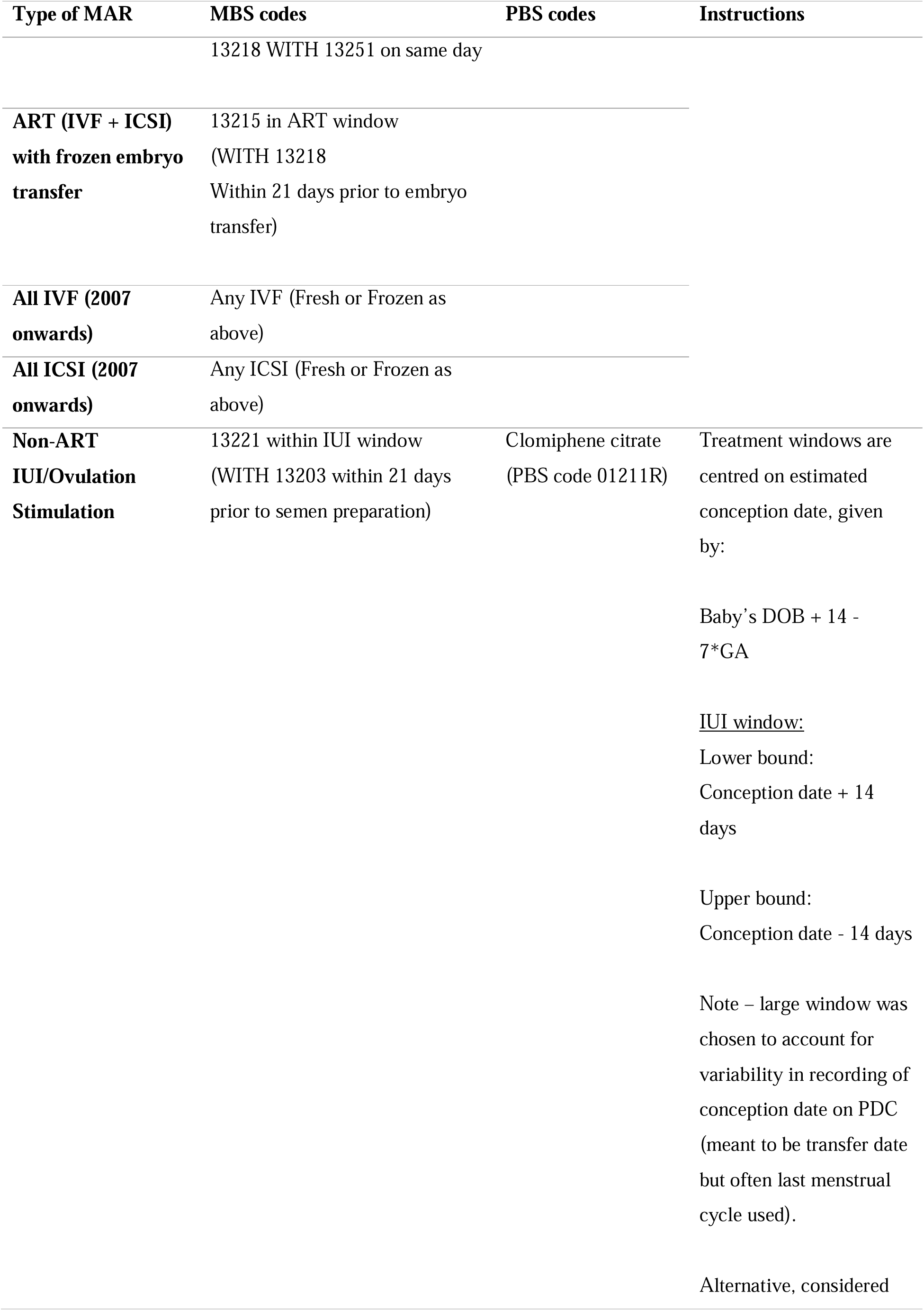
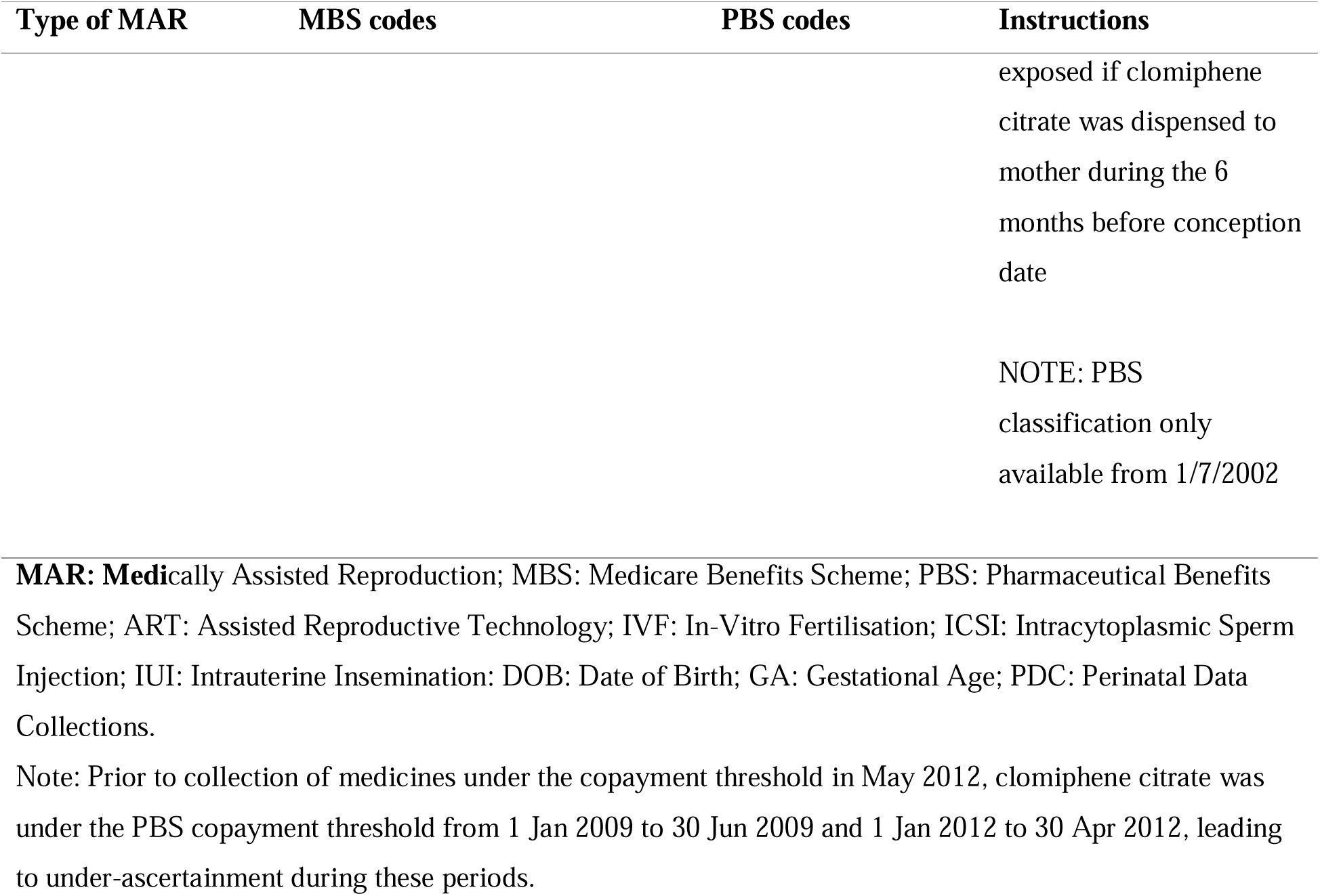
Definition of codes used to identify methods of conception and treatment windows for mother.

#### Other variable definitions

All variables were defined relative to study entry (date of birth). From the State/Territory Registries of Births, Deaths, and Marriages, we defined: calendar year of birth; child’s sex (male, female, or indeterminate); and maternal age at birth in years. From the State/Territory Perinatal Data Collections, we defined: the state of residence where the birth was registered; the approximate gestational age at birth (in weeks); maternal history of at least one prior pregnancy > 20 weeks gestation; maternal smoking during pregnancy; maternal history of diabetes (gestational or otherwise) prior to the child’s estimated conception date; and child’s sex where it was not recorded in the births registry. From the Medicare Enrolment File, we determined: maternal area-level indicator of residential remoteness as per the Accessibility/Remoteness Index of Australia Plus (ARIA+; classified as metropolitan, inner regional, remote/very remote) (Australian Centre for Housing Research, 2023); and maternal area-level indicator of socioeconomic disadvantage based on 2016 census data as per the Socio-Economic Indexes for Areas (SEIFA, defined in deciles) (Australian Bureau of Statistics, 2018). From the Australian Cancer Database we determined whether either registered parent had a notifiable cancer (any invasive cancer, in-situ breast cancer since 2002, in-situ melanoma since 2004) before the estimated conception date.

Some states (Victoria, 44.31%; Western Australia, 20.20%; South Australia, 21.35%) had a substantial proportion of missing maternal smoking history data, resulting in 16.74% of the cohort having no information. Consequently, smoking history was coded as either yes, no, or missing. Mothers who only had gestational diabetes on the current child’s birth record (but not previous children) were not considered to have a history of diabetes for that child, as conception (either natural or via MAR) occurred before the diagnosis of gestational diabetes. As NSW changed their ascertainment of diabetes in 2015, information for children born after this date was not available, and therefore all children born in NSW after 2015 were coded as “Missing” (6.82% of primary cohort).

For SIRs we calculated a time-varying variable of attained age (0-1, 1-4, 5-9, 10-14 years).

## Outcome variables

The AIHW linked all identified children to the Australian Cancer Database (1982-2019) and the National Death Index (1991-2019). As cancers and deaths are mandated reportable events in Australia, the data are population-based and high-quality (Australian Institute of Health and Welfare, 2024).

We defined childhood cancer based on the age at diagnosis, from birth until age 15. We examined all childhood cancers combined and the twelve cancer types classified by the International Classification of Childhood Cancer (third edition) (Steliarova-Foucher et al., 2017, Steliarova-Foucher et al., 2005). We also report a custom cancer category of embryonal cancers (Supplementary Table 2). All childhood cancers registered in the Australian Cancer Database are malignant.

## Statistical analysis

We calculated descriptive statistics for all variables for each type of MAR conception in the primary cohort. The mean, standard deviation, median, and interquartile range are reported for continuous variables. The total number and proportion are reported for discrete variables.

To describe the population incidence of childhood cancers in MAR-conceived singleton children, we calculated SIRs for each cancer for each type of MAR conception. SIRs were standardised by sex, age group, state, and calendar year. National cancer rates for singleton children were calculated using all MAR and naturally-conceived children. 95% confidence intervals were calculated using exact Poisson count tests. For conception via MAR, ART (any type), and OI/IUI, we also estimated SIRs for all cancers stratified by attained age, sex, whether the child was born preterm (<37 weeks gestation), and maternal age at birth (below or above the national median). We calculated the risk difference per 100,000 person-years for each cancer, based on the derived SIR (and 95% confidence intervals) and the expected number of cancers.

To estimate the association between MAR and cancer in singleton children, we conducted survival analysis with flexible parametric survival modelling using stabilised inverse probability weights. Inverse probability weights were calculated using logistic regression, with parental history of cancer, maternal age at birth, maternal history of diabetes, area-based socio-economic status, residential remoteness, previous pregnancy, and history of smoking included as confounders (balance tables reported in Supplementary Table 3; Love plots reported in Supplementary Figure 1). We used flexible parametric survival modelling (Royston and Lambert, 2011) with two splines in the baseline hazard to calculate hazard ratios comparing naturally-conceived children to children conceived via each type of MAR. To account for variance within siblings, we used robust clustering on the mother’s identifier. We calculated the cumulative marginal difference in incident cancers per 100,000 children by age 15 using the *standsurv* command (Lambert, 2021). This command calculates the expected extra number of cancers by age 15 for MAR-exposed children compared to naturally-conceived children, controlling for the mentioned confounding variables.

As there was evidence of a mismatch in mean maternal age at birth between naturally-conceived children and children conceived via ART after weighting (Supplementary Table 3, Supplementary Figure 1), for all survival models we conducted sensitivity analyses including maternal age at birth in the outcome model as well as in the calculation of inverse probability weights.

To maintain participant confidentiality and comply with data provider protocols, we are only permitted to report SIRs for cancers with six or more incident cases for children conceived via MAR. Similarly, when reporting results from the survival analysis, we report survival model statistics for cancers with six or more incident cases for children conceived via MAR and in the presentation of results we round the number of cancers reported to the closest ten.

Children were followed up from date of birth until either 31 December 2019, their 15^th^ birthday, or death, whichever occurred first. All analysis time was calculated in days since birth. For models splitting ICSI and IVF, only children born from 2007 and onwards were eligible for inclusion (including comparators), as ICSI and IVF could not be differentiated prior this time. For all other models we include all children. We report our findings in line with the STROBE reporting guidelines for observational cohort studies (Vandenbroucke et al., 2014).

## 4. Results

### Cohort demographics

In Table 2 (naturally-conceived, MAR, ART, OI/IUI) and Supplementary Table 4 (Other ART sub-groups) we show the demographic characteristics of singletons conceived using the MAR treatments and those conceived naturally. Overall, the annual numbers of singleton children conceived via MAR increased over the course of the study period. More children conceived via MAR were born to mothers living in major cities, and these mothers also tended to live in less disadvantaged areas compared to mothers of naturally-conceived children. Mothers of children conceived via MAR tended to be older (median age: 34 years) compared to mothers of naturally-conceived children (median age: 31 years). Children conceived via MAR were more likely to have had at least one parent with a cancer prior to conception (2.5%) compared to parents of naturally-conceived children (1.2%).

**Table 2:**
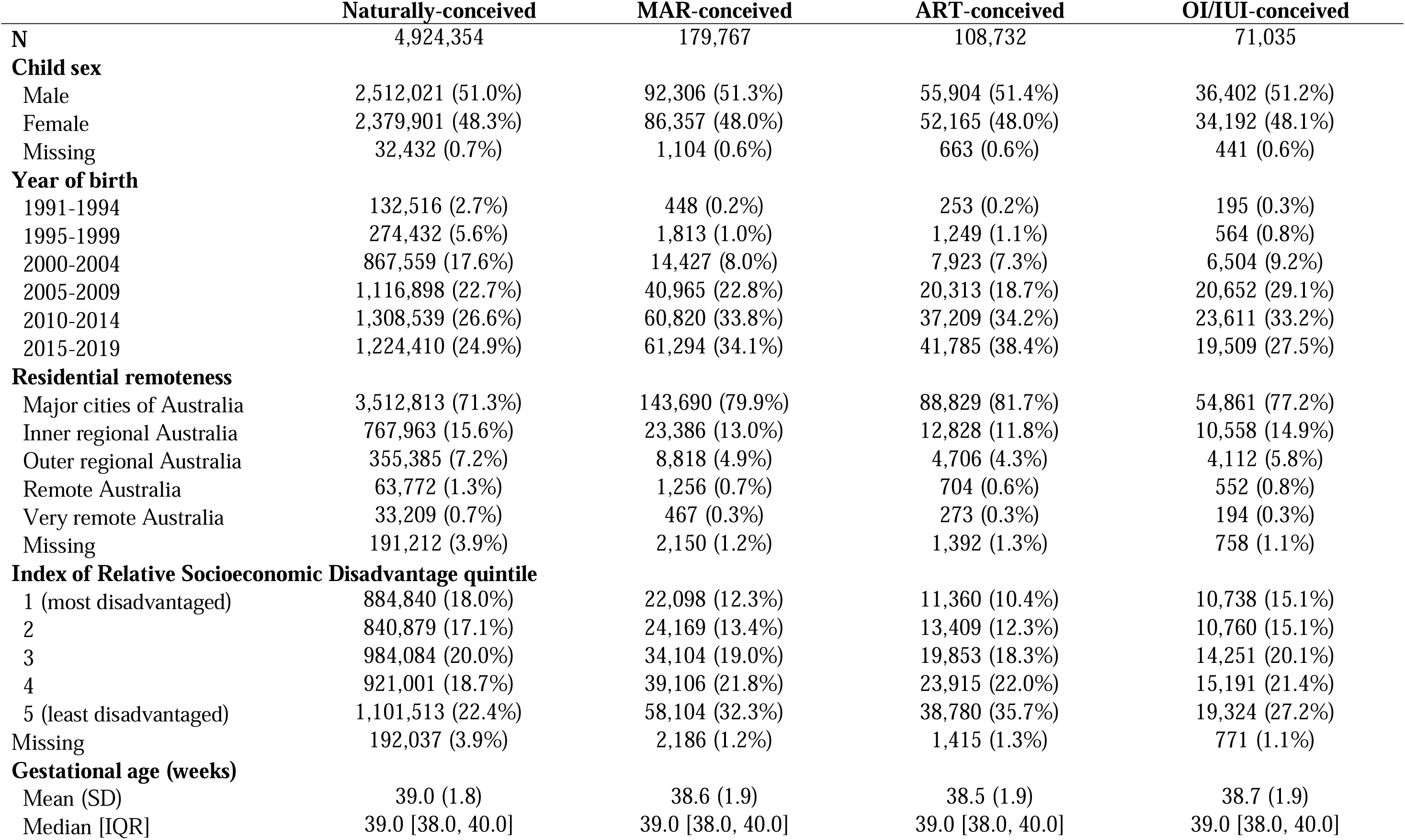
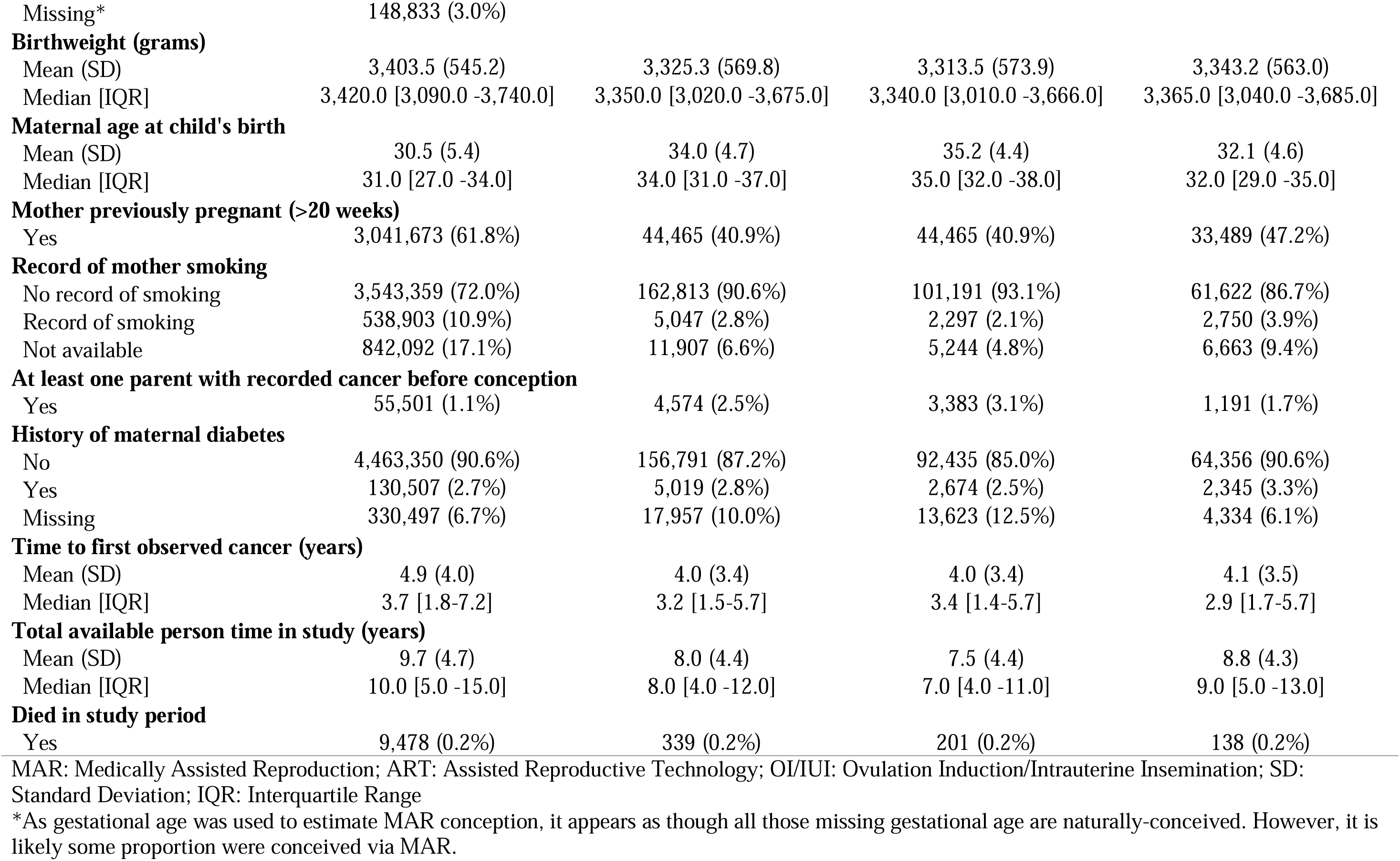
Cohort demographics table by treatment type for the primary cohort (MAR, ART, and OI/IUI)

|  | Naturally-conceived | MAR-conceived | ART-conceived | OI/UI-conceived |
| --- | --- | --- | --- | --- |
| **N** | 4,924,354 | 179,767 | 108,732 | 71,035 |
| **Child sex** |  |  |  |  |
| Male | 2,512,021 (51.0%) | 92,306 (51.3%) | 55,904 (51.4%) | 36,402 (51.2%) |
| Female | 2,379,901 (48.3%) | 86,357 (48.0%) | 52,165 (48.0%) | 34,192 (48.1%) |
| Missing | 32,432 (0.7%) | 1,104 (0.6%) | 663 (0.6%) | 441 (0.6%) |
| **Year of birth** |  |  |  |  |
| 1991-1994 | 132,516 (2.7%) | 448 (0.2%) | 253 (0.2%) | 195 (0.3%) |
| 1995-1999 | 274,432 (5.6%) | 1,813 (1.0%) | 1,249 (1.1%) | 564 (0.8%) |
| 2000-2004 | 867,559 (17.6%) | 14,427 (8.0%) | 7,923 (7.3%) | 6,504 (9.2%) |
| 2005-2009 | 1,116,898 (22.7%) | 40,965 (22.8%) | 20,313 (18.7%) | 20,652 (29.1%) |
| 2010-2014 | 1,308,539 (26.6%) | 60,820 (33.8%) | 37,209 (34.2%) | 23,611 (33.2%) |
| 2015-2019 | 1,224,410 (24.9%) | 61,294 (34.1%) | 41,785 (38.4%) | 19,509 (27.5%) |
| **Residential remoteness** |  |  |  |  |
| Major cities of Australia | 3,512,813 (71.3%) | 143,690 (79.9%) | 88,829 (81.7%) | 54,861 (77.2%) |
| Inner regional Australia | 767,963 (15.6%) | 23,386 (13.0%) | 12,828 (11.8%) | 10,558 (14.9%) |
| Outer regional Australia | 355,385 (7.2%) | 8,818 (4.9%) | 4,706 (4.3%) | 4,112 (5.8%) |
| Remote Australia | 63,772 (1.3%) | 1,256 (0.7%) | 704 (0.6%) | 552 (0.8%) |
| Very remote Australia | 33,209 (0.7%) | 467 (0.3%) | 273 (0.3%) | 194 (0.3%) |
| Missing | 191,212 (3.9%) | 2,150 (1.2%) | 1,392 (1.3%) | 758 (1.1%) |
| **Index of Relative Socioeconomic Disadvantage quintile** |  |  |  |  |
| 1 (most disadvantaged) | 884,840 (18.0%) | 22,098 (12.3%) | 11,360 (10.4%) | 10,738 (15.1%) |
| 2 | 840,879 (17.1%) | 24,169 (13.4%) | 13,409 (12.3%) | 10,760 (15.1%) |
| 3 | 984,084 (20.0%) | 34,104 (19.0%) | 19,853 (18.3%) | 14,251 (20.1%) |
| 4 | 921,001 (18.7%) | 39,106 (21.8%) | 23,915 (22.0%) | 15,191 (21.4%) |
| 5 (least disadvantaged) | 1,101,513 (22.4%) | 58,104 (32.3%) | 38,780 (35.7%) | 19,324 (27.2%) |
| Missing | 192,037 (3.9%) | 2,186 (1.2%) | 1,415 (1.3%) | 771 (1.1%) |
| **Gestational age (weeks)** |  |  |  |  |
| Mean (SD) | 39.0 (1.8) | 38.6 (1.9) | 38.5 (1.9) | 38.7 (1.9) |
| Median [IQR] | 39.0 [38.0, 40.0] | 39.0 [38.0, 40.0] | 39.0 [38.0, 40.0] | 39.0 [38.0, 40.0] |
| Missing* | 148,833 (3.0%) |  |  |  |
| **Birthweight (grams)** |  |  |  |  |
| Mean (SD) | 3,403.5 (545.2) | 3,325.3 (569.8) | 3,313.5 (573.9) | 3,343.2 (563.0) |
| Median [IQR] | 3,420.0 [3,090.0 -3,740.0] | 3,350.0 [3,020.0 -3,675.0] | 3,340.0 [3,010.0 -3,666.0] | 3,365.0 [3,040.0 -3,685.0] |
| **Maternal age at child's birth** |  |  |  |  |
| Mean (SD) | 30.5 (5.4) | 34.0 (4.7) | 35.2 (4.4) | 32.1 (4.6) |
| Median [IQR] | 31.0 [27.0 -34.0] | 34.0 [31.0 -37.0] | 35.0 [32.0 -38.0] | 32.0 [29.0 -35.0] |
| **Mother previously pregnant (>20 weeks)** |  |  |  |  |
| Yes | 3,041,673 (61.8%) | 44,465 (40.9%) | 44,465 (40.9%) | 33,489 (47.2%) |
| **Record of mother smoking** |  |  |  |  |
| No record of smoking | 3,543,359 (72.0%) | 162,813 (90.6%) | 101,191 (93.1%) | 61,622 (86.7%) |
| Record of smoking | 538,903 (10.9%) | 5,047 (2.8%) | 2,297 (2.1%) | 2,750 (3.9%) |
| Not available | 842,092 (17.1%) | 11,907 (6.6%) | 5,244 (4.8%) | 6,663 (9.4%) |
| **At least one parent with recorded cancer before conception** |  |  |  |  |
| Yes | 55,501 (1.1%) | 4,574 (2.5%) | 3,383 (3.1%) | 1,191 (1.7%) |
| **History of maternal diabetes** |  |  |  |  |
| No | 4,463,350 (90.6%) | 156,791 (87.2%) | 92,435 (85.0%) | 64,356 (90.6%) |
| Yes | 130,507 (2.7%) | 5,019 (2.8%) | 2,674 (2.5%) | 2,345 (3.3%) |
| Missing | 330,497 (6.7%) | 17,957 (10.0%) | 13,623 (12.5%) | 4,334 (6.1%) |
| **Time to first observed cancer (years)** |  |  |  |  |
| Mean (SD) | 4.9 (4.0) | 4.0 (3.4) | 4.0 (3.4) | 4.1 (3.5) |
| Median [IQR] | 3.7 [1.8-7.2] | 3.2 [1.5-5.7] | 3.4 [1.4-5.7] | 2.9 [1.7-5.7] |
| **Total available person time in study (years)** |  |  |  |  |
| Mean (SD) | 9.7 (4.7) | 8.0 (4.4) | 7.5 (4.4) | 8.8 (4.3) |
| Median [IQR] | 10.0 [5.0 -15.0] | 8.0 [4.0 -12.0] | 7.0 [4.0 -11.0] | 9.0 [5.0 -13.0] |
| **Died in study period** |  |  |  |  |
| Yes | 9,478 (0.2%) | 339 (0.2%) | 201 (0.2%) | 138 (0.2%) |

MAR: Medically Assisted Reproduction; ART: Assisted Reproductive Technology; OI/IUI: Ovulation Induction/Intrauterine Insemination; SD: Standard Deviation; IQR: Interquartile Range
\*As gestational age was used to estimate MAR conception, it appears as though all those missing gestational age are naturally-conceived. However, it is likely some proportion were conceived via MAR.

### Standardised incidence ratios

The SIRs for all cancers and treatment types are shown in Figure 2 (any MAR, ART, and OI/IUI) and Supplementary Figure 2 (IVF/ICSI and fresh/frozen embryo transfer). The all-cancer stratified SIRs are presented in Supplementary Figures 3-5. There was no increased all-cancer incidence in singleton children following conception by MAR treatment at the 95% confidence level.

**Figure 2:**
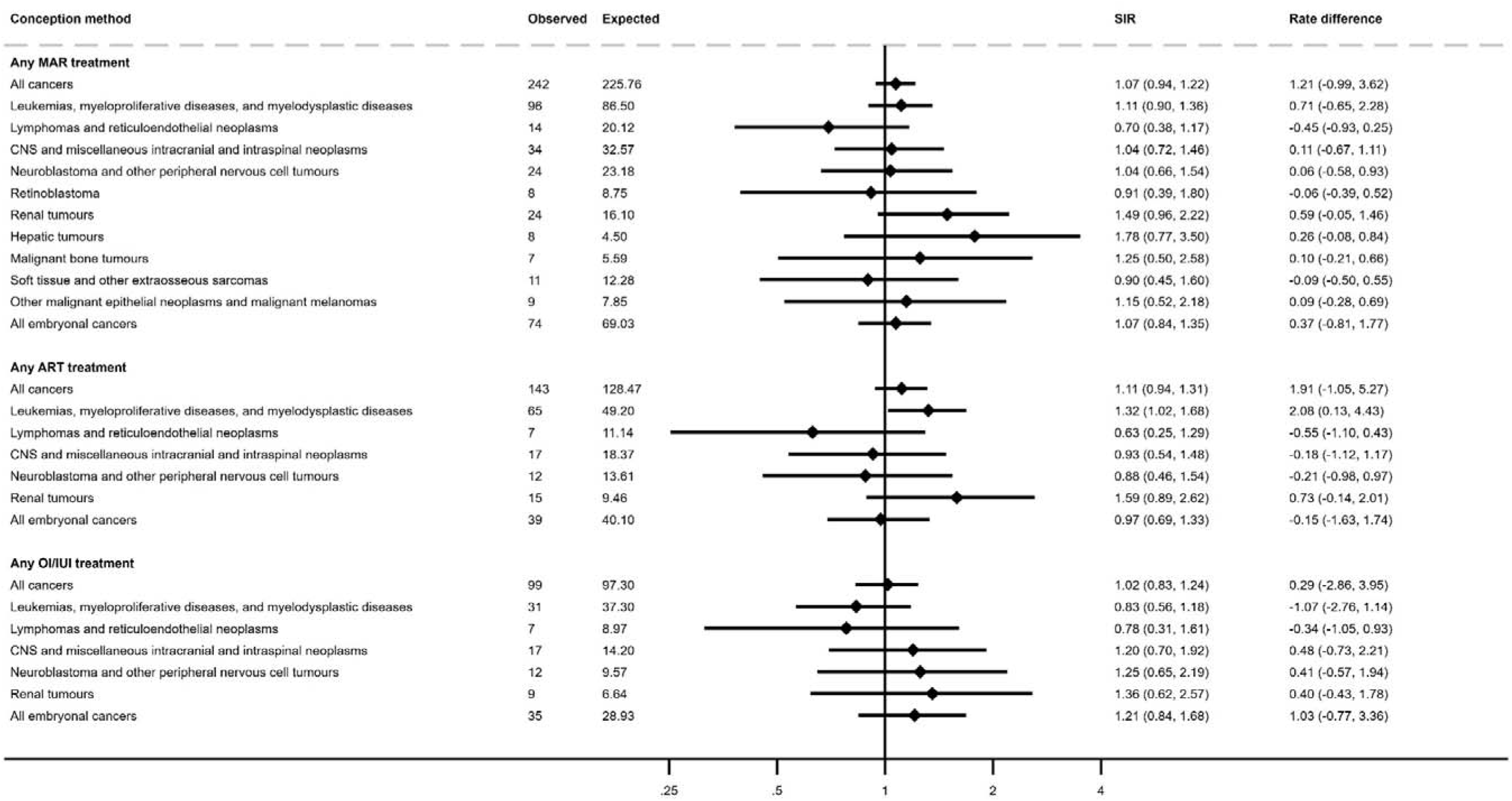
Standardised incidence ratios and risk differences for childhood cancers for children conceived using any medically assisted reproduction (MAR), any assisted reproductive treatment (ART) and any ovulation induction/intrauterine insemination (OI/IUI).

For specific cancer groups, only four exhibited an elevated incidence in children conceived by MAR compared to the age and sex-matched general population of children. They were leukemias, myeloproliferative diseases, and myelodysplastic diseases for children conceived via ART (SIR: 1.32, 95% confidence interval: 1.02-1.68) and for children conceived via IVF treatment (1.54, 1.01-2.26), and renal tumours for children conceived via frozen ART treatment (2.52, 1.09-4.97) and children conceived via IVF treatment (2.37, 1.02-4.67). These cancers had a risk difference of fewer than six extra cancers per 100,000 person-years (at the top 95% confidence interval).

Stratification of all-cancer SIRs by child age, sex at birth, whether born at term or before, and maternal age (above or below median) showed no meaningful differences for MAR-conceived and naturally-conceived children (Supplementary Figures 3-5).

### Survival analysis

Figure 3 (MAR, ART, and OI/IUI) and Supplementary Figure 6 (IVF/ICSI and fresh/frozen embryo transfer) show the results from survival modelling. Due to issues with model convergence, models estimating the risk of renal tumours and hepatic tumours were fitted without any splines in the baseline hazard. There was no evidence any of the cancers tested showed an elevated risk following MAR, ART, or OI/IUI after controlling for the listed confounding variables. When analysing ART sub-groups, there was an increased risk of leukemias, myeloproliferative diseases, and myelodysplastic diseases following IVF treatment (hazard ratio: 1.77, 95% confidence interval: 1.06-2.95), but children conceived via frozen transfer ICSI treatment had a reduced risk of this cancer group compared to naturally-conceived children (hazard ratio: 0.33, 95% confidence interval: 0.15-0.76). Sensitivity analyses with maternal age at birth also included in the outcome model (Supplementary Figures 7 and 8) suggested no meaningful change in results from inverse probability of treatment weighting alone.

**Figure 3:**
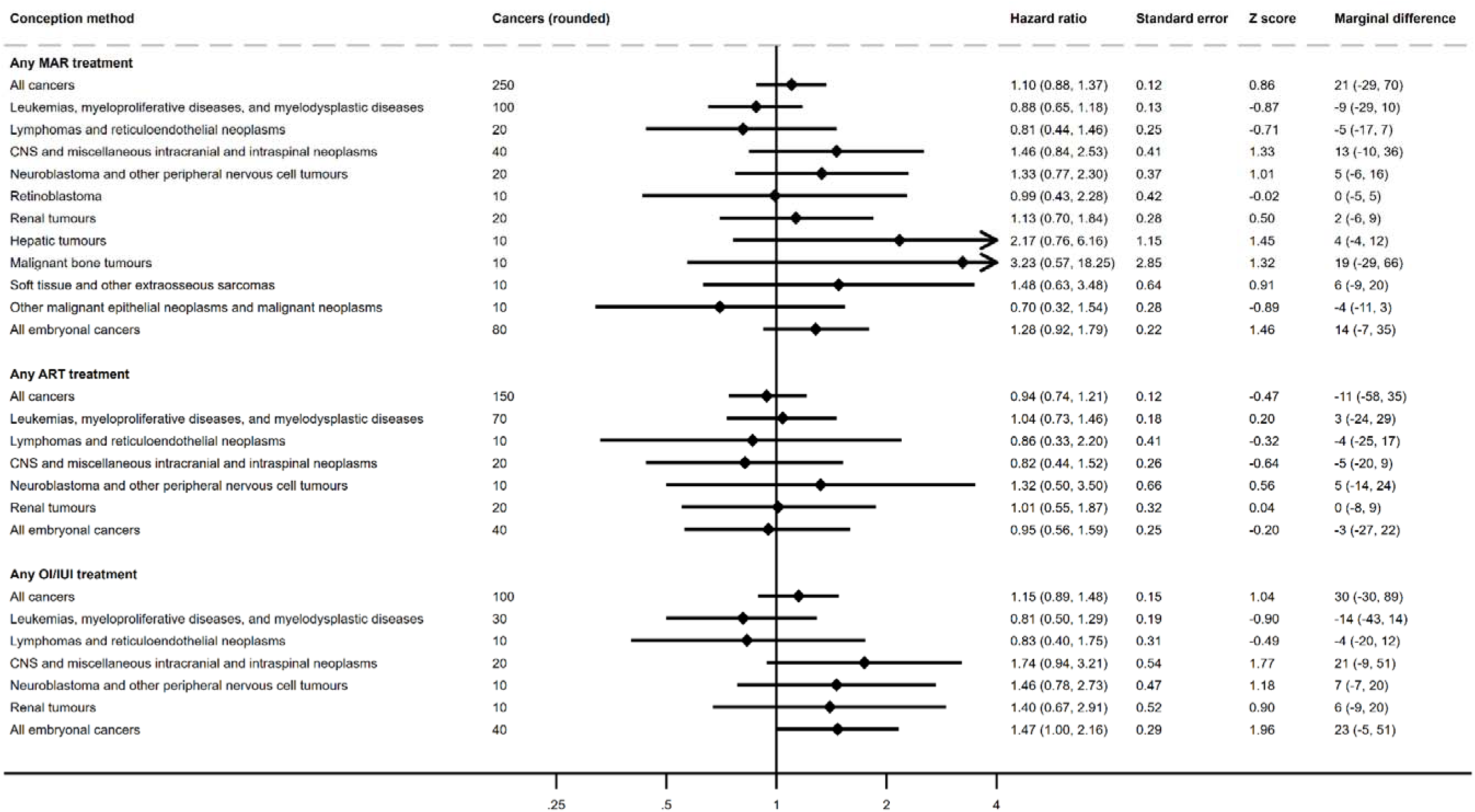
Hazard ratios and cumulative marginal cancer difference for childhood cancers for children conceived using any medically assisted reproduction (MAR), any assisted reproductive treatment (ART) and any ovulation induction/intrauterine insemination (OI/IUI).

## 5. Discussion and recommendations

Our study described the population incidence and estimated the association with childhood cancer for singletons conceived via MAR and born in Australia between 1991 and 2019.

Compared to the age-, sex-, calendar year-and state-matched general population, singletons conceived via ART may have a slightly elevated incidence of leukemias, myeloproliferative diseases, and myelodysplastic diseases, as well as renal tumours, following IVF conception or frozen transfer ART conception. However, we found limited evidence MAR conception itself (either via ART or OI/IUI methods) is associated with an increased risk of childhood cancer after controlling for key confounding variables. These findings provide reassurance that singletons conceived via MAR are not at an increased risk of cancer compared to their naturally-conceived counterparts.

While we noted in our study the leukemias, myeloproliferative diseases, and myelodysplastic diseases group occurred at elevated incidence in MAR-conceived singletons compared to the general population (as per the SIR), an elevated risk was generally not evident after controlling for additional confounding factors, specifically parental history of cancer, maternal age at birth, maternal history of diabetes, area-based socio-economic status, residential remoteness, previous pregnancy, and history of smoking.

The exception to this finding was for those conceived via IVF treatment, where the risk for leukemias, myeloproliferative diseases, and myelodysplastic diseases remained elevated after controlling for the listed confounders. Though it is possible this finding represents a real risk of IVF conception on this group of malignancies, we note two competing explanations. The first is uncontrolled confounding by female factor infertility. Previous work has suggested an effect (HR: 1.30) of female factor infertility on childhood leukemia (Hargreave et al., 2013). Given those conceived via MAR would overwhelmingly be born to couples experiencing infertility, and we could not control for cause of infertility in our study, it is possible this association drove the effect we observed on leukemias, myeloproliferative diseases, and myelodysplastic diseases for children conceived via IVF. This explanation may also account for why we observed no association for children conceived via ICSI, given ICSI is generally targeted towards those experiencing male factor infertility (though the application of ICSI in Australia varies considerably, McPherson et al. (2021)). Alternatively, given the proximity of the lower-bound confidence interval to 1, and the number of comparisons performed, the observed relationship between IVF conception and leukemias, myeloproliferative diseases, and myelodysplastic diseases may reflect a type I error (‘false positive’). For the same reason, we recommend caution about the decreased risk of leukemias, myeloproliferative diseases, and myelodysplastic diseases for those conceived via ICSI with frozen embryo treatment.

We also saw weak evidence for an increased incidence of renal tumours following ART conception with frozen embryo transfer, and IVF conception by any embryo transfer method compared to the general population. Again, this effect was not observed when controlling for confounding. Wilm’s tumour is the most prevalent paediatric renal cancer, accounting for over 90% of renal tumours in children aged 1-7 years (Nakata et al., 2020). It is possible Wilm’s tumours specifically may be increased for children conceived via ART, driven by an increased risk of imprinting disorders (Charlton et al., 2017, Kagami et al., 2025). However, if this were the case we would expect this finding to be robust to confounder adjustment.

There is evidence pre-term birth is associated with both an increased risk of Wilm’s tumours (Chu et al., 2010) and advanced maternal age at birth (Esposito et al., 2022). However, evidence maternal age itself is related to an increase in Wilm’s tumours is mixed (Chu, et al., 2010). Our results suggest further investigation of the maternal factors that influence the development of Wilm’s tumours may be warranted to unpack any potential relationship with conception via MAR.

Finally, it is uncertain whether hepatic tumours may be elevated for singletons conceived via MAR. Though hepatic tumours showed the highest point estimate of all the cancer subgroups examined, low precision and concerns over model estimation leaves us unable to determine whether this reflects a real association. One population-based UK study found an SIR of 3.27 (1.20-7.12) for hepatic tumours in children conceived via autologous (non-donor) ART (Williams et al., 2013), though this study did not find an increased incidence of childhood cancers overall, and did not control for variables other than age, sex and calendar year. More research with larger cohorts is needed to determine whether there is an association between conception via MAR and hepatic tumours.

Overall, our findings align with two recent population-based studies that show limited or no effect of conception via MAR on the development of cancer (Oakley, et al., 2025, Rios, et al., 2024). Furthermore, our study shows no strong evidence those conceived via frozen-embryo transfer are at any overall greater risk of childhood cancer, at odds with two recent, but partly overlapping Nordic population-based studies (Hargreave, et al., 2019, Sargisian, et al., 2022), or that children conceived via ICSI are at greater risk of cancer as reported in a Dutch study (Spaan, et al., 2023).

Inconsistencies between our findings and those of prior population-based studies is perhaps not unexpected given the small sample sizes for cancer groups, and chance of Type 1 errors. Larger studies where cohorts are combined across countries and/or federated meta-analysis are performed using such cohorts will allow more precise estimates to any risk to be determined. Furthermore, freezing techniques, culture media, freezing solutions, and transfer fluids have changed over the course of our study period and continue to differ across countries, which could contribute to inconsistent findings between countries.

### Strengths and limitations

Better understanding of any links between ART and subsequent childhood cancer risk is vital. For individuals/young families, this knowledge may inform their fertility-related care choices. At a broader policy and health-system level, this knowledge is critical to informing safe, optimal approaches to fertility-related healthcare. Our study advances knowledge on this question by examining over 5 million children using a population-based study design over an extended time in a country where MAR treatments are common. Cancer incidence is examined with respect to MAR type and cancer type, with the latter based on the International Classification of Childhood Cancer groupings and a custom group of embryonal cancers based on prior evidence (Spector, et al., 2019).

As with other studies, we were challenged by the rarity of childhood cancer, as well as limited follow-up time for some MAR treatments. This low incidence led to low precision in the estimates of the impact of MAR conception on individual cancer types. It is also likely low incidence contributed to inconsistencies between the marginal predicted number of cancers and hazard ratios around the extreme ends of the confidence interval, as both calculations are sensitive to small cell sizes. We also were unable to control for cause of parental infertility or subfertility, or cancer predisposition syndrome. Maternal infertility has been associated with a modestly increased risk (18%) of childhood cancers (primarily leukemias and cancers of the endocrine glands) compared to women without infertility issues (Hargreave, et al., 2013), though it is unclear whether particular causes of infertility may be more consequential for childhood cancers than others.

We were restricted to using an estimation of MAR conception based on gestational age and mother’s treatment history. However, around 3% of children did not have a recorded gestational age at birth, and we were therefore unable to determine whether they were conceived via MAR. We assume these values were missing at random given our adjustment set (particularly socioeconomic status and rurality), though we cannot rule out the potential for bias from the exclusion of these children. Additionally, clomiphene citrate use was not ascertained from 1 Jan 2009 to 30 Jun 2009 and 1 Jan 2012 to 30 Apr 2012 due to how medicines were reimbursed through the PBS during this time. As such, some misclassification during this period for conception assisted by clomiphene citrate use is also likely.

Finally, we did not capture incident benign or borderline tumours. Though most cancer groupings for the International Classification of Childhood Cancer only consider malignant tumours, two groupings (Central Nervous System and Miscellaneous Intracranial and Intraspinal Neoplasms; Germ Cell Tumours, Trophoblastic Tumors, and Neoplasms of the Gonads) also include benign and borderline tumours. Though we have no strong reason to believe the exclusion of benign and borderline tumours would change our effect estimates, readers should be aware of their exclusion when interpreting our findings.

## Conclusions

Our study converges with international literature in showing no evidence of an increased overall incidence or association with childhood cancer for singletons conceived via MAR, regardless of whether conception is by ART or OI/IUI. Some specific cancers may occur at increased incidence compared to the general population, although this can likely be attributed to differences in maternal characteristics, and the total number of extra expected cancers is small (at most fewer than 6 per 100,000 person-years). Our findings are generally concordant with recent population-based evidence, with no consistency between studies in relation to excess risk of specific cancer groups or types. Overall, we believe these results provide reassurance to healthcare practitioners delivering MAR, those who have undertaken or are considering MAR, and those born to MAR treatments on the safety of the treatments. We recommend future work focus on identifying ways to increase precision in estimates, either through federated meta-analysis or combining person-level data across jurisdictions.

## 6. Author’s roles

Authors ARW, CMV, SO, GMC, and CV contributed to the study design. Authors CMV, GMC, and CV contributed to acquisition of data. Authors ARW, CMV, SO, and CV contributed to analysis of data. All authors contributed to the interpretation of data. All authors contributed to either drafting the article or revising it critically for important intellectual content, reviewed the final version of the article for submission, and agree to be accountable for all aspects of the work in ensuring that questions related to the accuracy or integrity of any part of the work are appropriately investigated and resolved.

## Supporting information

STROBE

## 7. Acknowledgements

The authors are grateful for the perspectives and advice offered by Maree Pickens, consumer advocate, in the interpretation of the findings. We also acknowledge and thank the staff at the Australian Institute of Health and Welfare Data Linkage Unit, the Data Access and Assurance Unit, Data Services Branch, Queensland Health, the NSW Centre for Health Record Linkage, the Tasmanian Data Linkage Unit, the Centre for Victorian Data Linkage, and the Linkage, Data Outputs and ISPD Client Services teams at Western Australia Data Linkage Services for supporting the project and undertaking data linkage. We thank the Australian Institute of Health and Welfare and the population-based cancer registries of New South Wales (NSW), Victoria, Queensland, Western Australia, South Australia, Tasmania, the Australian Capital Territory (ACT) and the Northern Territory for the provision of data from the Australian Cancer Database. We also thank the following data custodians for providing the datasets used for this project: Australian Department of Health and Aged Care (Medicare Benefits Schedule, Pharmaceutical Benefits Scheme); Australian Institute of Health and Welfare (National Death Index); the NSW Ministry of Health, the Department of Health Victoria, the Data Services Branch Queensland Health, the Department of Health Western Australia, Preventive Health South Australia, the Department of Health Tasmania, HealthInfo ACT (perinatal data); and the jurisdictional Registries of Births, Deaths and Marriages (birth data). We are grateful to the Victorian Consultative Council on Obstetric and Paediatric Mortality and Morbidity (CCOPMM) for providing access to the data used for this project and for the assistance of the staff at Safer Care Victoria. The conclusions, findings, opinions and views or recommendations expressed in this paper are strictly those of the authors. They do not necessarily reflect those of CCOPMM.

## 9. Data availability

The analyses were based on data from different registries and administrative datasets (Australian Department of Health and Aged Care, State and Territory Health Departments, and the Australian Institute of Health and Welfare). The data can be made available on request to each of the data custodians after ethical approval from the relevant Human Research Ethics Committees. The statistical analysis plan is available at https://osf.io/rk9n6 and analysis code at https://osf.io/nqkc2/.

