## Supplementary for "Childhood cancer in singletons conceived via medically assisted reproduction in Australia: a population-based cohort study"

[Supplementary Figures 1 A-I: Love plots for balance of confounding variables before and after weighting. These map onto confounding variables as follows: Mothers_age: Maternal age at birth (numeric); smoke: History of smoking (1 = No, 2 = Yes, 3 = Missing); prev_preg: Any history of previous pregnancy (1 = Yes, 2 = No); remoteness: Remoteness of residence (1-4 in order of increasing remoteness, 5 = Missing); IRSD: Index of Relative Socioeconomic Disadvantage of Area of Residence Decile (1-10 in order of decreasing disadvantage, 11 = Missing); parent_cancer: History of cancer in either parent (1 = No, 2 = Yes); diabetes_history: Maternal history of diabetes (1 = No, 2 = Yes, 3 = Missing).	19](#_Toc226621157)

### 1. Supplementary Tables

#### Supplementary Table 1: Description of datasets used in linkage

| Dataset | Timeframe | Original purpose and setting | Use in cohort |
| --- | --- | --- | --- |
| National datasets |  |  |  |
| Medicare Enrolment File (MEF)  (Australian National Audit Office, 2024) | 1 Jan 1991 - 31 Dec 2019 | The MEF records information on individuals enrolled in Medicare (the Australian public health insurance program). It records names and addresses of those enrolled. Enrolment occurs either shortly after birth, or, in the case of immigrants, when they become eligible. | Identifies key demographic information at the time of birth of the child from a linked mother/father/child triad, as well as identifying mothers and fathers who had not used Medicare services and those who had immigrated. |
| Medicare Benefits Schedule (MBS)  (Australian Institute of Health and Welfare, 2024b) | 1 Jan 1991 - 31 Dec 2019 | The MBS data collection records all health service claims made through the Medicare Benefits Schedule. These claims are provided by healthcare providers who are reimbursed through the Schedule. | Used to identify a MAR treatment for the mother around the time of conception, and therefore determine conception via MAR for the child (in conjunction with the PBS).  Researchers only received MAR relevant items (See Table 1) |
| Pharmaceutical Benefits Scheme (PBS)  (Australian Institute of Health and Welfare, 2024d) | 1 Jul 2002 - 31 Dec 2019 | The PBS data collection records all health service claims made through the Pharmaceutical Benefits Scheme (the Australian public medicines reimbursement program). These claims are provided by pharmacists who are reimbursed through the Schedule. | Used to identify dispensations of clomiphene citrate to the mother around the critical time for conception, and therefore determine assisted conception for the child (in conjunction with the MBS - See Table 1). |
| National Death Index (NDI)  (Australian Institute of Health and Welfare, 2024c) | 1 Jan 1991 - 31 Dec 2019 | Registry recording all deaths that occur in Australia. Reporting of all deaths occurring in Australia is mandated and recorded in this data collection. Obtained from the Australian Institute of Health and Welfare. | Used to identify time of the death for children for the purposes of censoring in SIR calculations and survival modelling. |
| Australian Cancer Database (ACD)  (Australian Institute of Health and Welfare, 2024a) | 1 Jan 1982 - 31 Dec 2019 | National cancer registry recording all notifiable cancers reported to Australia’s state-based cancer registries. Reporting of all notifiable cancers is mandatory. | Used to identify time, topography, and morphology of a given cancer diagnosis. |
| State datasets |  |  |  |
| Perinatal Data Collections (PDCs) for NSW, ACT, QLD, VIC, WA*, and TAS | **NSW:** 1 Jan 1994 –  30 Jun 2019  **ACT:** 1 Jan 1997 –  1 Dec 2017  **QLD:** 1 Jan 2007 - 31 Dec 2019  **VIC:** 1 Jan 1999 - 20 Oct 2022  **SA:** 1 Jan 1991 - 31 Dec 2019  **WA:** 1 Jan 1991 - 31 Dec 2020 **TAS:** 1 Jan 2005 - 31 Dec 2019 | The PDCs are State-based data collections recording information on pregnancies beyond 20 weeks gestation (or above 400 grams in weight in the case of Victoria).  All States include date of birth, status of the birth (live or still), plurality, parity, complications of pregnancy, and other relevant data items, though the precise nature of recording and data items differ between States. | Used in combination with the RBDM to identify births in Australia, and relevant information about pregnancy for use in SIR calculations and survival modelling. |
| Registry of Births, Deaths, and Marriages (RBDM) for NSW, ACT, QLD, VIC, WA, and TAS | **NSW:** 1 Jan 1994 –  30 Jun 2019  **ACT:** 1 Jan 1997 –  1 Dec 2017  **QLD:** 1 Jan 2007 - 31 Dec 2018  **VIC:** 1 Jan 1999 - 20 Oct 2022  **SA:** 1 Jan 1991 - 31 Dec 2019  **WA:** 1 Jan 1991 - 31 Dec 2020 **TAS:** 1 Jan 2005 - 31 Dec 2019 | The RBDM records information on all births (including information on mothers and fathers) recorded in the relevant State or Territory. | Used in combination with the PDC datasets to identify births in Australia, create mother/father/child triads, and identify certain information for SIR calculations and survival modelling. |
| NSW=New South Wales; ACT=Australian Capital Territory; QLD=Queensland; VIC=Victoria; SA=South Australia; WA=Western Australia; TAS=Tasmania.  *Known as the “Western Australia Midwife Notification System”. | | | |

#### Supplementary Table 2: Custom embryonal cancer group

| **ICD-O-3 topography  (3 characters unless otherwise specified)** | **ICD-O-3.1 morphology (behaviour code 3)** | **Subgroup** |
| --- | --- | --- |
| All | 9490, 9500 | Neuroblastoma |
| All | 9510-9514 | Retinoblastoma |
| All | 8959, 8960 | Nephroblastoma (Wilms tumor) |
| C64.9 | 8963, 8964 |  |
| All | 8970 | Hepatoblastoma |
| All | 8900–8905, 8910, 8912, 8920, 8991 | Embryonal rhabdomyosarcoma |
| All | 8972, 8973 | Pulmonary and pleuropulmonary blastoma |
| All | 9470–9472, 9474, 9480 | Medulloblastoma |
| All | 9473 | PNET(Primitive neuroectodermal tumor) |
| C70.0–C72.9 | 9501–9504 | Medulloepithelioma |
| All | 9508 | Atypical teratoid rhabdoid tumours |
| ICD-O-3: International Classification of Diseases for Oncology (3^rd^ edition) | | |

#### Supplementary Table 3: Balance of confounding variables before and after weighting

|  |  | **Unweighted** | | | | | **Weighted** | | | | |
| --- | --- | --- | --- | --- | --- | --- | --- | --- | --- | --- | --- |
|  |  | **Exposed** | | **Unexposed** | |  | **Exposed** | | **Unexposed** | |  |
| **Exposure** | **Variable** | **Mean** | **SD** | **Mean** | **SD** | **SMD** | **Mean** | **SD** | **Mean** | **SD** | **SMD** |
| MAR | mothers_age | 33.99 | 4.70 | 30.50 | 5.39 | 0.69 | 31.14 | 4.82 | 30.63 | 5.42 | 0.10 |
|  | smoke_1 | 0.91 | 0.29 | 0.74 | 0.44 | 0.44 | 0.76 | 0.43 | 0.75 | 0.43 | 0.02 |
|  | smoke_2 | 0.03 | 0.17 | 0.11 | 0.32 | 0.34 | 0.11 | 0.31 | 0.11 | 0.31 | 0.00 |
|  | smoke_3 | 0.07 | 0.25 | 0.15 | 0.35 | 0.26 | 0.13 | 0.34 | 0.14 | 0.35 | 0.03 |
|  | prev_preg_1 | 0.57 | 0.50 | 0.38 | 0.49 | 0.38 | 0.37 | 0.48 | 0.39 | 0.49 | 0.04 |
|  | prev_preg_2 | 0.43 | 0.50 | 0.62 | 0.49 | 0.38 | 0.63 | 0.48 | 0.61 | 0.49 | 0.04 |
|  | remoteness_1 | 0.80 | 0.40 | 0.72 | 0.45 | 0.20 | 0.72 | 0.45 | 0.72 | 0.45 | 0.00 |
|  | remoteness_2 | 0.13 | 0.34 | 0.15 | 0.36 | 0.07 | 0.15 | 0.36 | 0.15 | 0.36 | 0.00 |
|  | remoteness_3 | 0.05 | 0.22 | 0.07 | 0.26 | 0.10 | 0.07 | 0.26 | 0.07 | 0.26 | 0.00 |
|  | remoteness_4 | 0.01 | 0.10 | 0.02 | 0.14 | 0.09 | 0.02 | 0.13 | 0.02 | 0.14 | 0.02 |
|  | remoteness_5 | 0.01 | 0.11 | 0.04 | 0.19 | 0.17 | 0.04 | 0.19 | 0.04 | 0.19 | 0.00 |
|  | IRSD_1 | 0.06 | 0.24 | 0.10 | 0.29 | 0.13 | 0.10 | 0.30 | 0.10 | 0.29 | 0.01 |
|  | IRSD_2 | 0.06 | 0.24 | 0.08 | 0.28 | 0.09 | 0.08 | 0.28 | 0.08 | 0.28 | 0.00 |
|  | IRSD_3 | 0.06 | 0.23 | 0.07 | 0.26 | 0.06 | 0.07 | 0.26 | 0.07 | 0.26 | 0.00 |
|  | IRSD_4 | 0.08 | 0.27 | 0.10 | 0.30 | 0.07 | 0.09 | 0.29 | 0.10 | 0.30 | 0.02 |
|  | IRSD_5 | 0.09 | 0.28 | 0.10 | 0.30 | 0.04 | 0.10 | 0.30 | 0.10 | 0.30 | 0.00 |
|  | IRSD_6 | 0.10 | 0.30 | 0.10 | 0.30 | 0.01 | 0.10 | 0.30 | 0.10 | 0.30 | 0.00 |
|  | IRSD_7 | 0.09 | 0.29 | 0.08 | 0.28 | 0.04 | 0.09 | 0.28 | 0.08 | 0.28 | 0.01 |
|  | IRSD_8 | 0.12 | 0.33 | 0.10 | 0.30 | 0.06 | 0.10 | 0.30 | 0.10 | 0.31 | 0.00 |
|  | IRSD_9 | 0.15 | 0.36 | 0.12 | 0.32 | 0.11 | 0.12 | 0.32 | 0.12 | 0.32 | 0.00 |
|  | IRSD_10 | 0.17 | 0.38 | 0.11 | 0.31 | 0.18 | 0.11 | 0.31 | 0.11 | 0.31 | 0.00 |
|  | IRSD_11 | 0.01 | 0.11 | 0.04 | 0.19 | 0.17 | 0.04 | 0.19 | 0.04 | 0.19 | 0.00 |
|  | parent_cancer_1 | 0.97 | 0.16 | 0.99 | 0.11 | 0.11 | 0.99 | 0.11 | 0.99 | 0.11 | 0.01 |
|  | parent_cancer_2 | 0.03 | 0.16 | 0.01 | 0.11 | 0.11 | 0.01 | 0.11 | 0.01 | 0.11 | 0.01 |
|  | diabetes_history_1 | 0.87 | 0.33 | 0.90 | 0.29 | 0.10 | 0.90 | 0.30 | 0.90 | 0.30 | 0.01 |
|  | diabetes_history_2 | 0.03 | 0.16 | 0.03 | 0.16 | 0.01 | 0.03 | 0.17 | 0.03 | 0.16 | 0.02 |
|  | diabetes_history_3 | 0.10 | 0.30 | 0.07 | 0.25 | 0.11 | 0.07 | 0.26 | 0.07 | 0.26 | 0.00 |
| ART | mothers_age | 35.20 | 4.38 | 30.50 | 5.39 | 0.96 | 32.03 | 4.39 | 30.60 | 5.43 | 0.29 |
|  | smoke_1 | 0.93 | 0.25 | 0.74 | 0.44 | 0.53 | 0.78 | 0.41 | 0.75 | 0.44 | 0.08 |
|  | smoke_2 | 0.02 | 0.14 | 0.11 | 0.32 | 0.37 | 0.09 | 0.29 | 0.11 | 0.31 | 0.06 |
|  | smoke_3 | 0.05 | 0.21 | 0.15 | 0.35 | 0.33 | 0.13 | 0.33 | 0.14 | 0.35 | 0.05 |
|  | prev_preg_1 | 0.59 | 0.49 | 0.38 | 0.49 | 0.43 | 0.38 | 0.49 | 0.39 | 0.49 | 0.01 |
|  | prev_preg_2 | 0.41 | 0.49 | 0.62 | 0.49 | 0.43 | 0.62 | 0.49 | 0.61 | 0.49 | 0.01 |
|  | remoteness_1 | 0.82 | 0.39 | 0.72 | 0.45 | 0.24 | 0.73 | 0.45 | 0.72 | 0.45 | 0.02 |
|  | remoteness_2 | 0.12 | 0.32 | 0.15 | 0.36 | 0.11 | 0.15 | 0.35 | 0.15 | 0.36 | 0.02 |
|  | remoteness_3 | 0.04 | 0.20 | 0.07 | 0.26 | 0.13 | 0.07 | 0.25 | 0.07 | 0.26 | 0.02 |
|  | remoteness_4 | 0.01 | 0.09 | 0.02 | 0.14 | 0.09 | 0.02 | 0.13 | 0.02 | 0.14 | 0.02 |
|  | remoteness_5 | 0.01 | 0.11 | 0.04 | 0.19 | 0.16 | 0.04 | 0.20 | 0.04 | 0.19 | 0.03 |
|  | IRSD_1 | 0.05 | 0.22 | 0.10 | 0.29 | 0.18 | 0.09 | 0.29 | 0.10 | 0.29 | 0.01 |
|  | IRSD_2 | 0.05 | 0.23 | 0.08 | 0.28 | 0.12 | 0.08 | 0.27 | 0.08 | 0.28 | 0.02 |
|  | IRSD_3 | 0.05 | 0.22 | 0.07 | 0.26 | 0.09 | 0.07 | 0.25 | 0.07 | 0.26 | 0.02 |
|  | IRSD_4 | 0.07 | 0.26 | 0.10 | 0.30 | 0.09 | 0.09 | 0.28 | 0.10 | 0.30 | 0.03 |
|  | IRSD_5 | 0.08 | 0.28 | 0.10 | 0.30 | 0.06 | 0.09 | 0.29 | 0.10 | 0.30 | 0.02 |
|  | IRSD_6 | 0.10 | 0.30 | 0.10 | 0.30 | 0.01 | 0.10 | 0.30 | 0.10 | 0.30 | 0.00 |
|  | IRSD_7 | 0.09 | 0.29 | 0.08 | 0.28 | 0.03 | 0.09 | 0.28 | 0.08 | 0.28 | 0.01 |
|  | IRSD_8 | 0.13 | 0.33 | 0.10 | 0.30 | 0.08 | 0.11 | 0.31 | 0.10 | 0.31 | 0.01 |
|  | IRSD_9 | 0.16 | 0.37 | 0.12 | 0.32 | 0.13 | 0.12 | 0.33 | 0.12 | 0.32 | 0.02 |
|  | IRSD_10 | 0.20 | 0.40 | 0.11 | 0.31 | 0.25 | 0.12 | 0.33 | 0.11 | 0.31 | 0.03 |
|  | IRSD_11 | 0.01 | 0.11 | 0.04 | 0.19 | 0.16 | 0.04 | 0.20 | 0.04 | 0.19 | 0.03 |
|  | parent_cancer_1 | 0.97 | 0.17 | 0.99 | 0.11 | 0.14 | 0.98 | 0.12 | 0.99 | 0.11 | 0.03 |
|  | parent_cancer_2 | 0.03 | 0.17 | 0.01 | 0.11 | 0.14 | 0.02 | 0.12 | 0.01 | 0.11 | 0.03 |
|  | diabetes_history_1 | 0.85 | 0.36 | 0.90 | 0.29 | 0.17 | 0.90 | 0.31 | 0.90 | 0.30 | 0.02 |
|  | diabetes_history_2 | 0.02 | 0.15 | 0.03 | 0.16 | 0.01 | 0.03 | 0.17 | 0.03 | 0.16 | 0.01 |
|  | diabetes_history_3 | 0.13 | 0.33 | 0.07 | 0.25 | 0.19 | 0.08 | 0.26 | 0.07 | 0.26 | 0.02 |
| OI/IUI | mothers_age | 32.13 | 4.58 | 30.50 | 5.39 | 0.33 | 31.01 | 4.65 | 30.52 | 5.39 | 0.10 |
|  | smoke_1 | 0.87 | 0.34 | 0.74 | 0.44 | 0.32 | 0.76 | 0.43 | 0.74 | 0.44 | 0.03 |
|  | smoke_2 | 0.04 | 0.19 | 0.11 | 0.32 | 0.28 | 0.11 | 0.31 | 0.11 | 0.32 | 0.01 |
|  | smoke_3 | 0.09 | 0.29 | 0.15 | 0.35 | 0.16 | 0.13 | 0.34 | 0.15 | 0.35 | 0.03 |
|  | prev_preg_1 | 0.53 | 0.50 | 0.38 | 0.49 | 0.30 | 0.38 | 0.48 | 0.38 | 0.49 | 0.02 |
|  | prev_preg_2 | 0.47 | 0.50 | 0.62 | 0.49 | 0.30 | 0.62 | 0.48 | 0.62 | 0.49 | 0.02 |
|  | remoteness_1 | 0.77 | 0.42 | 0.72 | 0.45 | 0.13 | 0.72 | 0.45 | 0.72 | 0.45 | 0.01 |
|  | remoteness_2 | 0.15 | 0.36 | 0.15 | 0.36 | 0.02 | 0.15 | 0.36 | 0.15 | 0.36 | 0.00 |
|  | remoteness_3 | 0.06 | 0.23 | 0.07 | 0.26 | 0.06 | 0.07 | 0.26 | 0.07 | 0.26 | 0.00 |
|  | remoteness_4 | 0.01 | 0.10 | 0.02 | 0.14 | 0.08 | 0.02 | 0.13 | 0.02 | 0.14 | 0.01 |
|  | remoteness_5 | 0.01 | 0.10 | 0.04 | 0.19 | 0.18 | 0.04 | 0.19 | 0.04 | 0.19 | 0.01 |
|  | IRSD_1 | 0.08 | 0.27 | 0.10 | 0.29 | 0.06 | 0.10 | 0.29 | 0.10 | 0.29 | 0.00 |
|  | IRSD_2 | 0.07 | 0.26 | 0.08 | 0.28 | 0.05 | 0.08 | 0.28 | 0.08 | 0.28 | 0.00 |
|  | IRSD_3 | 0.07 | 0.25 | 0.07 | 0.26 | 0.03 | 0.07 | 0.26 | 0.07 | 0.26 | 0.00 |
|  | IRSD_4 | 0.09 | 0.28 | 0.10 | 0.30 | 0.04 | 0.10 | 0.29 | 0.10 | 0.30 | 0.01 |
|  | IRSD_5 | 0.09 | 0.29 | 0.10 | 0.30 | 0.03 | 0.10 | 0.30 | 0.10 | 0.30 | 0.00 |
|  | IRSD_6 | 0.11 | 0.31 | 0.10 | 0.30 | 0.03 | 0.10 | 0.30 | 0.10 | 0.30 | 0.00 |
|  | IRSD_7 | 0.10 | 0.30 | 0.08 | 0.28 | 0.05 | 0.08 | 0.28 | 0.08 | 0.28 | 0.00 |
|  | IRSD_8 | 0.12 | 0.32 | 0.10 | 0.30 | 0.04 | 0.10 | 0.30 | 0.10 | 0.30 | 0.00 |
|  | IRSD_9 | 0.14 | 0.34 | 0.12 | 0.32 | 0.07 | 0.12 | 0.32 | 0.12 | 0.32 | 0.00 |
|  | IRSD_10 | 0.13 | 0.34 | 0.11 | 0.31 | 0.08 | 0.11 | 0.31 | 0.11 | 0.31 | 0.01 |
|  | IRSD_11 | 0.01 | 0.10 | 0.04 | 0.19 | 0.18 | 0.04 | 0.19 | 0.04 | 0.19 | 0.01 |
|  | parent_cancer_1 | 0.98 | 0.13 | 0.99 | 0.11 | 0.05 | 0.99 | 0.11 | 0.99 | 0.11 | 0.01 |
|  | parent_cancer_2 | 0.02 | 0.13 | 0.01 | 0.11 | 0.05 | 0.01 | 0.11 | 0.01 | 0.11 | 0.01 |
|  | diabetes_history_1 | 0.91 | 0.29 | 0.90 | 0.29 | 0.01 | 0.90 | 0.30 | 0.90 | 0.29 | 0.02 |
|  | diabetes_history_2 | 0.03 | 0.18 | 0.03 | 0.16 | 0.04 | 0.03 | 0.17 | 0.03 | 0.16 | 0.01 |
|  | diabetes_history_3 | 0.06 | 0.24 | 0.07 | 0.25 | 0.03 | 0.07 | 0.26 | 0.07 | 0.25 | 0.02 |
| Any IVF | mothers_age | 35.44 | 4.39 | 30.73 | 5.40 | 0.96 | 32.26 | 4.33 | 30.79 | 5.43 | 0.30 |
|  | smoke_1 | 0.96 | 0.20 | 0.86 | 0.35 | 0.36 | 0.87 | 0.33 | 0.86 | 0.35 | 0.04 |
|  | smoke_2 | 0.02 | 0.14 | 0.11 | 0.32 | 0.38 | 0.10 | 0.30 | 0.11 | 0.31 | 0.04 |
|  | smoke_3 | 0.02 | 0.15 | 0.03 | 0.17 | 0.05 | 0.03 | 0.17 | 0.03 | 0.17 | 0.01 |
|  | prev_preg_1 | 0.56 | 0.50 | 0.38 | 0.49 | 0.35 | 0.37 | 0.48 | 0.39 | 0.49 | 0.04 |
|  | prev_preg_2 | 0.44 | 0.50 | 0.62 | 0.49 | 0.35 | 0.63 | 0.48 | 0.61 | 0.49 | 0.04 |
|  | remoteness_1 | 0.83 | 0.38 | 0.73 | 0.45 | 0.24 | 0.74 | 0.44 | 0.73 | 0.45 | 0.04 |
|  | remoteness_2 | 0.11 | 0.32 | 0.16 | 0.37 | 0.15 | 0.15 | 0.36 | 0.16 | 0.37 | 0.03 |
|  | remoteness_3 | 0.05 | 0.21 | 0.08 | 0.26 | 0.12 | 0.07 | 0.26 | 0.08 | 0.26 | 0.00 |
|  | remoteness_4 | 0.01 | 0.10 | 0.02 | 0.13 | 0.07 | 0.02 | 0.13 | 0.02 | 0.13 | 0.01 |
|  | remoteness_5 | 0.00 | 0.07 | 0.02 | 0.12 | 0.11 | 0.01 | 0.11 | 0.02 | 0.12 | 0.03 |
|  | IRSD_1 | 0.05 | 0.22 | 0.10 | 0.30 | 0.18 | 0.09 | 0.29 | 0.10 | 0.30 | 0.02 |
|  | IRSD_2 | 0.05 | 0.22 | 0.09 | 0.28 | 0.13 | 0.08 | 0.27 | 0.08 | 0.28 | 0.02 |
|  | IRSD_3 | 0.05 | 0.21 | 0.07 | 0.26 | 0.11 | 0.07 | 0.25 | 0.07 | 0.26 | 0.01 |
|  | IRSD_4 | 0.07 | 0.26 | 0.10 | 0.30 | 0.09 | 0.09 | 0.29 | 0.10 | 0.30 | 0.01 |
|  | IRSD_5 | 0.08 | 0.28 | 0.10 | 0.30 | 0.06 | 0.10 | 0.30 | 0.10 | 0.30 | 0.01 |
|  | IRSD_6 | 0.10 | 0.30 | 0.11 | 0.31 | 0.02 | 0.10 | 0.30 | 0.11 | 0.31 | 0.01 |
|  | IRSD_7 | 0.09 | 0.29 | 0.09 | 0.28 | 0.03 | 0.09 | 0.29 | 0.09 | 0.28 | 0.01 |
|  | IRSD_8 | 0.13 | 0.34 | 0.11 | 0.31 | 0.06 | 0.12 | 0.32 | 0.11 | 0.31 | 0.02 |
|  | IRSD_9 | 0.16 | 0.36 | 0.12 | 0.32 | 0.12 | 0.13 | 0.33 | 0.12 | 0.32 | 0.04 |
|  | IRSD_10 | 0.20 | 0.40 | 0.11 | 0.31 | 0.27 | 0.12 | 0.32 | 0.11 | 0.31 | 0.03 |
|  | IRSD_11 | 0.00 | 0.07 | 0.02 | 0.13 | 0.11 | 0.01 | 0.11 | 0.02 | 0.12 | 0.03 |
|  | parent_cancer_1 | 0.98 | 0.15 | 0.99 | 0.11 | 0.09 | 0.98 | 0.12 | 0.99 | 0.11 | 0.02 |
|  | parent_cancer_2 | 0.02 | 0.15 | 0.01 | 0.11 | 0.09 | 0.02 | 0.12 | 0.01 | 0.11 | 0.02 |
|  | diabetes_history_1 | 0.81 | 0.39 | 0.87 | 0.34 | 0.15 | 0.86 | 0.34 | 0.86 | 0.34 | 0.00 |
|  | diabetes_history_2 | 0.03 | 0.16 | 0.03 | 0.18 | 0.04 | 0.03 | 0.18 | 0.03 | 0.18 | 0.00 |
|  | diabetes_history_3 | 0.17 | 0.37 | 0.10 | 0.30 | 0.19 | 0.10 | 0.30 | 0.10 | 0.30 | 0.00 |
| Any ICSI | mothers_age | 35.16 | 4.36 | 30.73 | 5.40 | 0.90 | 32.13 | 4.45 | 30.80 | 5.43 | 0.27 |
|  | smoke_1 | 0.97 | 0.18 | 0.86 | 0.35 | 0.40 | 0.88 | 0.32 | 0.86 | 0.35 | 0.08 |
|  | smoke_2 | 0.02 | 0.14 | 0.11 | 0.32 | 0.38 | 0.09 | 0.28 | 0.11 | 0.31 | 0.08 |
|  | smoke_3 | 0.01 | 0.12 | 0.03 | 0.17 | 0.12 | 0.03 | 0.17 | 0.03 | 0.17 | 0.01 |
|  | prev_preg_1 | 0.60 | 0.49 | 0.38 | 0.49 | 0.43 | 0.38 | 0.48 | 0.39 | 0.49 | 0.03 |
|  | prev_preg_2 | 0.40 | 0.49 | 0.62 | 0.49 | 0.43 | 0.62 | 0.48 | 0.61 | 0.49 | 0.03 |
|  | remoteness_1 | 0.82 | 0.38 | 0.73 | 0.45 | 0.23 | 0.75 | 0.44 | 0.73 | 0.45 | 0.04 |
|  | remoteness_2 | 0.12 | 0.33 | 0.16 | 0.37 | 0.12 | 0.16 | 0.36 | 0.16 | 0.37 | 0.02 |
|  | remoteness_3 | 0.04 | 0.20 | 0.08 | 0.26 | 0.14 | 0.07 | 0.25 | 0.08 | 0.26 | 0.02 |
|  | remoteness_4 | 0.01 | 0.09 | 0.02 | 0.13 | 0.08 | 0.02 | 0.13 | 0.02 | 0.13 | 0.01 |
|  | remoteness_5 | 0.00 | 0.06 | 0.02 | 0.12 | 0.12 | 0.01 | 0.11 | 0.02 | 0.12 | 0.03 |
|  | IRSD_1 | 0.05 | 0.22 | 0.10 | 0.30 | 0.18 | 0.09 | 0.29 | 0.10 | 0.29 | 0.01 |
|  | IRSD_2 | 0.05 | 0.23 | 0.09 | 0.28 | 0.12 | 0.08 | 0.27 | 0.08 | 0.28 | 0.01 |
|  | IRSD_3 | 0.05 | 0.22 | 0.07 | 0.26 | 0.09 | 0.07 | 0.26 | 0.07 | 0.26 | 0.01 |
|  | IRSD_4 | 0.07 | 0.26 | 0.10 | 0.30 | 0.10 | 0.09 | 0.28 | 0.10 | 0.30 | 0.04 |
|  | IRSD_5 | 0.09 | 0.28 | 0.10 | 0.30 | 0.05 | 0.10 | 0.30 | 0.10 | 0.30 | 0.01 |
|  | IRSD_6 | 0.10 | 0.30 | 0.11 | 0.31 | 0.02 | 0.11 | 0.31 | 0.11 | 0.31 | 0.00 |
|  | IRSD_7 | 0.10 | 0.29 | 0.09 | 0.28 | 0.03 | 0.09 | 0.29 | 0.09 | 0.28 | 0.01 |
|  | IRSD_8 | 0.13 | 0.34 | 0.11 | 0.31 | 0.07 | 0.12 | 0.32 | 0.11 | 0.31 | 0.02 |
|  | IRSD_9 | 0.17 | 0.37 | 0.12 | 0.32 | 0.15 | 0.12 | 0.33 | 0.12 | 0.32 | 0.02 |
|  | IRSD_10 | 0.18 | 0.39 | 0.11 | 0.31 | 0.22 | 0.12 | 0.32 | 0.11 | 0.31 | 0.04 |
|  | IRSD_11 | 0.00 | 0.06 | 0.02 | 0.13 | 0.12 | 0.01 | 0.11 | 0.02 | 0.12 | 0.03 |
|  | parent_cancer_1 | 0.96 | 0.19 | 0.99 | 0.11 | 0.16 | 0.98 | 0.13 | 0.99 | 0.11 | 0.04 |
|  | parent_cancer_2 | 0.04 | 0.19 | 0.01 | 0.11 | 0.16 | 0.02 | 0.13 | 0.01 | 0.11 | 0.04 |
|  | diabetes_history_1 | 0.84 | 0.36 | 0.87 | 0.34 | 0.06 | 0.86 | 0.35 | 0.86 | 0.34 | 0.03 |
|  | diabetes_history_2 | 0.03 | 0.16 | 0.03 | 0.18 | 0.04 | 0.03 | 0.18 | 0.03 | 0.18 | 0.01 |
|  | diabetes_history_3 | 0.13 | 0.34 | 0.10 | 0.30 | 0.08 | 0.11 | 0.31 | 0.10 | 0.30 | 0.02 |
| Fresh ART | mothers_age | 34.82 | 4.27 | 30.50 | 5.39 | 0.89 | 32.00 | 4.39 | 30.56 | 5.41 | 0.29 |
|  | smoke_1 | 0.91 | 0.28 | 0.74 | 0.44 | 0.47 | 0.78 | 0.42 | 0.74 | 0.44 | 0.08 |
|  | smoke_2 | 0.02 | 0.15 | 0.11 | 0.32 | 0.36 | 0.09 | 0.29 | 0.11 | 0.31 | 0.06 |
|  | smoke_3 | 0.06 | 0.24 | 0.15 | 0.35 | 0.27 | 0.13 | 0.33 | 0.14 | 0.35 | 0.05 |
|  | prev_preg_1 | 0.66 | 0.47 | 0.38 | 0.49 | 0.57 | 0.37 | 0.48 | 0.39 | 0.49 | 0.03 |
|  | prev_preg_2 | 0.34 | 0.47 | 0.62 | 0.49 | 0.57 | 0.63 | 0.48 | 0.61 | 0.49 | 0.03 |
|  | remoteness_1 | 0.81 | 0.39 | 0.72 | 0.45 | 0.23 | 0.73 | 0.44 | 0.72 | 0.45 | 0.03 |
|  | remoteness_2 | 0.12 | 0.32 | 0.15 | 0.36 | 0.11 | 0.14 | 0.35 | 0.15 | 0.36 | 0.03 |
|  | remoteness_3 | 0.04 | 0.21 | 0.07 | 0.26 | 0.12 | 0.07 | 0.25 | 0.07 | 0.26 | 0.02 |
|  | remoteness_4 | 0.01 | 0.10 | 0.02 | 0.14 | 0.09 | 0.02 | 0.13 | 0.02 | 0.14 | 0.02 |
|  | remoteness_5 | 0.02 | 0.12 | 0.04 | 0.19 | 0.14 | 0.04 | 0.20 | 0.04 | 0.19 | 0.03 |
|  | IRSD_1 | 0.05 | 0.22 | 0.10 | 0.29 | 0.17 | 0.09 | 0.28 | 0.10 | 0.29 | 0.02 |
|  | IRSD_2 | 0.06 | 0.23 | 0.08 | 0.28 | 0.11 | 0.08 | 0.26 | 0.08 | 0.28 | 0.03 |
|  | IRSD_3 | 0.05 | 0.22 | 0.07 | 0.26 | 0.08 | 0.07 | 0.25 | 0.07 | 0.26 | 0.01 |
|  | IRSD_4 | 0.07 | 0.26 | 0.10 | 0.30 | 0.09 | 0.09 | 0.29 | 0.10 | 0.30 | 0.03 |
|  | IRSD_5 | 0.09 | 0.28 | 0.10 | 0.30 | 0.05 | 0.09 | 0.29 | 0.10 | 0.30 | 0.02 |
|  | IRSD_6 | 0.10 | 0.30 | 0.10 | 0.30 | 0.01 | 0.10 | 0.30 | 0.10 | 0.30 | 0.01 |
|  | IRSD_7 | 0.09 | 0.29 | 0.08 | 0.28 | 0.03 | 0.09 | 0.28 | 0.08 | 0.28 | 0.02 |
|  | IRSD_8 | 0.13 | 0.33 | 0.10 | 0.30 | 0.07 | 0.11 | 0.31 | 0.10 | 0.31 | 0.02 |
|  | IRSD_9 | 0.16 | 0.36 | 0.12 | 0.32 | 0.12 | 0.12 | 0.33 | 0.12 | 0.32 | 0.03 |
|  | IRSD_10 | 0.19 | 0.39 | 0.11 | 0.31 | 0.24 | 0.12 | 0.32 | 0.11 | 0.31 | 0.03 |
|  | IRSD_11 | 0.02 | 0.12 | 0.04 | 0.19 | 0.14 | 0.04 | 0.20 | 0.04 | 0.19 | 0.03 |
|  | parent_cancer_1 | 0.97 | 0.17 | 0.99 | 0.11 | 0.13 | 0.98 | 0.13 | 0.99 | 0.11 | 0.04 |
|  | parent_cancer_2 | 0.03 | 0.17 | 0.01 | 0.11 | 0.13 | 0.02 | 0.13 | 0.01 | 0.11 | 0.04 |
|  | diabetes_history_1 | 0.88 | 0.32 | 0.90 | 0.29 | 0.06 | 0.90 | 0.30 | 0.90 | 0.29 | 0.02 |
|  | diabetes_history_2 | 0.02 | 0.14 | 0.03 | 0.16 | 0.04 | 0.03 | 0.17 | 0.03 | 0.16 | 0.01 |
|  | diabetes_history_3 | 0.10 | 0.29 | 0.07 | 0.25 | 0.10 | 0.07 | 0.26 | 0.07 | 0.25 | 0.02 |
| Frozen ART | mothers_age | 35.83 | 4.48 | 30.50 | 5.39 | 1.08 | 32.15 | 4.28 | 30.54 | 5.41 | 0.33 |
|  | smoke_1 | 0.96 | 0.20 | 0.74 | 0.44 | 0.64 | 0.78 | 0.42 | 0.74 | 0.44 | 0.07 |
|  | smoke_2 | 0.02 | 0.13 | 0.11 | 0.32 | 0.40 | 0.10 | 0.30 | 0.11 | 0.32 | 0.05 |
|  | smoke_3 | 0.02 | 0.15 | 0.15 | 0.35 | 0.45 | 0.13 | 0.33 | 0.14 | 0.35 | 0.05 |
|  | prev_preg_1 | 0.48 | 0.50 | 0.38 | 0.49 | 0.20 | 0.37 | 0.48 | 0.38 | 0.49 | 0.03 |
|  | prev_preg_2 | 0.52 | 0.50 | 0.62 | 0.49 | 0.20 | 0.63 | 0.48 | 0.62 | 0.49 | 0.03 |
|  | remoteness_1 | 0.82 | 0.38 | 0.72 | 0.45 | 0.25 | 0.71 | 0.45 | 0.72 | 0.45 | 0.01 |
|  | remoteness_2 | 0.12 | 0.33 | 0.15 | 0.36 | 0.10 | 0.16 | 0.36 | 0.15 | 0.36 | 0.01 |
|  | remoteness_3 | 0.04 | 0.20 | 0.07 | 0.26 | 0.13 | 0.07 | 0.26 | 0.07 | 0.26 | 0.01 |
|  | remoteness_4 | 0.01 | 0.09 | 0.02 | 0.14 | 0.10 | 0.02 | 0.13 | 0.02 | 0.14 | 0.03 |
|  | remoteness_5 | 0.01 | 0.09 | 0.04 | 0.19 | 0.20 | 0.04 | 0.20 | 0.04 | 0.19 | 0.02 |
|  | IRSD_1 | 0.05 | 0.21 | 0.10 | 0.29 | 0.19 | 0.10 | 0.30 | 0.10 | 0.29 | 0.01 |
|  | IRSD_2 | 0.05 | 0.22 | 0.08 | 0.28 | 0.13 | 0.08 | 0.28 | 0.08 | 0.28 | 0.00 |
|  | IRSD_3 | 0.05 | 0.21 | 0.07 | 0.26 | 0.11 | 0.07 | 0.25 | 0.07 | 0.26 | 0.01 |
|  | IRSD_4 | 0.07 | 0.26 | 0.10 | 0.30 | 0.10 | 0.09 | 0.28 | 0.10 | 0.30 | 0.03 |
|  | IRSD_5 | 0.08 | 0.27 | 0.10 | 0.30 | 0.07 | 0.09 | 0.29 | 0.10 | 0.30 | 0.03 |
|  | IRSD_6 | 0.10 | 0.30 | 0.10 | 0.30 | 0.00 | 0.10 | 0.30 | 0.10 | 0.30 | 0.00 |
|  | IRSD_7 | 0.09 | 0.29 | 0.08 | 0.28 | 0.04 | 0.08 | 0.28 | 0.08 | 0.28 | 0.00 |
|  | IRSD_8 | 0.13 | 0.34 | 0.10 | 0.30 | 0.08 | 0.11 | 0.31 | 0.10 | 0.30 | 0.01 |
|  | IRSD_9 | 0.17 | 0.37 | 0.12 | 0.32 | 0.15 | 0.12 | 0.32 | 0.12 | 0.32 | 0.01 |
|  | IRSD_10 | 0.20 | 0.40 | 0.11 | 0.31 | 0.26 | 0.12 | 0.32 | 0.11 | 0.31 | 0.03 |
|  | IRSD_11 | 0.01 | 0.09 | 0.04 | 0.19 | 0.20 | 0.04 | 0.20 | 0.04 | 0.19 | 0.02 |
|  | parent_cancer_1 | 0.97 | 0.18 | 0.99 | 0.11 | 0.16 | 0.99 | 0.12 | 0.99 | 0.11 | 0.03 |
|  | parent_cancer_2 | 0.03 | 0.18 | 0.01 | 0.11 | 0.16 | 0.01 | 0.12 | 0.01 | 0.11 | 0.03 |
|  | diabetes_history_1 | 0.79 | 0.40 | 0.90 | 0.29 | 0.31 | 0.89 | 0.31 | 0.90 | 0.30 | 0.03 |
|  | diabetes_history_2 | 0.03 | 0.18 | 0.03 | 0.16 | 0.03 | 0.03 | 0.17 | 0.03 | 0.16 | 0.02 |
|  | diabetes_history_3 | 0.17 | 0.38 | 0.07 | 0.25 | 0.32 | 0.08 | 0.26 | 0.07 | 0.26 | 0.02 |
| Fresh IVF | mothers_age | 34.93 | 4.20 | 30.73 | 5.40 | 0.87 | 32.22 | 4.33 | 30.76 | 5.41 | 0.30 |
|  | smoke_1 | 0.95 | 0.22 | 0.86 | 0.35 | 0.32 | 0.87 | 0.34 | 0.86 | 0.35 | 0.04 |
|  | smoke_2 | 0.02 | 0.14 | 0.11 | 0.32 | 0.38 | 0.10 | 0.30 | 0.11 | 0.32 | 0.03 |
|  | smoke_3 | 0.03 | 0.17 | 0.03 | 0.17 | 0.01 | 0.03 | 0.17 | 0.03 | 0.17 | 0.02 |
|  | prev_preg_1 | 0.63 | 0.48 | 0.38 | 0.49 | 0.52 | 0.36 | 0.48 | 0.39 | 0.49 | 0.05 |
|  | prev_preg_2 | 0.37 | 0.48 | 0.62 | 0.49 | 0.52 | 0.64 | 0.48 | 0.61 | 0.49 | 0.05 |
|  | remoteness_1 | 0.83 | 0.38 | 0.73 | 0.45 | 0.25 | 0.74 | 0.44 | 0.73 | 0.45 | 0.04 |
|  | remoteness_2 | 0.11 | 0.31 | 0.16 | 0.37 | 0.16 | 0.15 | 0.36 | 0.16 | 0.37 | 0.04 |
|  | remoteness_3 | 0.05 | 0.21 | 0.08 | 0.26 | 0.12 | 0.07 | 0.26 | 0.08 | 0.26 | 0.00 |
|  | remoteness_4 | 0.01 | 0.10 | 0.02 | 0.13 | 0.07 | 0.02 | 0.13 | 0.02 | 0.13 | 0.01 |
|  | remoteness_5 | 0.00 | 0.07 | 0.02 | 0.12 | 0.11 | 0.01 | 0.11 | 0.02 | 0.12 | 0.02 |
|  | IRSD_1 | 0.05 | 0.22 | 0.10 | 0.30 | 0.17 | 0.09 | 0.28 | 0.10 | 0.30 | 0.04 |
|  | IRSD_2 | 0.05 | 0.23 | 0.09 | 0.28 | 0.12 | 0.07 | 0.26 | 0.09 | 0.28 | 0.04 |
|  | IRSD_3 | 0.05 | 0.22 | 0.07 | 0.26 | 0.10 | 0.07 | 0.26 | 0.07 | 0.26 | 0.00 |
|  | IRSD_4 | 0.07 | 0.26 | 0.10 | 0.30 | 0.09 | 0.10 | 0.30 | 0.10 | 0.30 | 0.01 |
|  | IRSD_5 | 0.09 | 0.28 | 0.10 | 0.30 | 0.05 | 0.10 | 0.30 | 0.10 | 0.30 | 0.00 |
|  | IRSD_6 | 0.10 | 0.30 | 0.11 | 0.31 | 0.02 | 0.10 | 0.30 | 0.11 | 0.31 | 0.02 |
|  | IRSD_7 | 0.09 | 0.29 | 0.09 | 0.28 | 0.02 | 0.09 | 0.29 | 0.09 | 0.28 | 0.01 |
|  | IRSD_8 | 0.13 | 0.33 | 0.11 | 0.31 | 0.06 | 0.12 | 0.32 | 0.11 | 0.31 | 0.02 |
|  | IRSD_9 | 0.15 | 0.36 | 0.12 | 0.32 | 0.12 | 0.13 | 0.34 | 0.12 | 0.32 | 0.05 |
|  | IRSD_10 | 0.20 | 0.40 | 0.11 | 0.31 | 0.26 | 0.12 | 0.32 | 0.11 | 0.31 | 0.03 |
|  | IRSD_11 | 0.00 | 0.07 | 0.02 | 0.13 | 0.11 | 0.01 | 0.11 | 0.02 | 0.12 | 0.02 |
|  | parent_cancer_1 | 0.98 | 0.14 | 0.99 | 0.11 | 0.07 | 0.98 | 0.12 | 0.99 | 0.11 | 0.03 |
|  | parent_cancer_2 | 0.02 | 0.14 | 0.01 | 0.11 | 0.07 | 0.02 | 0.12 | 0.01 | 0.11 | 0.03 |
|  | diabetes_history_1 | 0.84 | 0.36 | 0.87 | 0.34 | 0.06 | 0.87 | 0.34 | 0.86 | 0.34 | 0.01 |
|  | diabetes_history_2 | 0.02 | 0.14 | 0.03 | 0.18 | 0.08 | 0.03 | 0.18 | 0.03 | 0.18 | 0.01 |
|  | diabetes_history_3 | 0.14 | 0.34 | 0.10 | 0.30 | 0.10 | 0.10 | 0.30 | 0.10 | 0.30 | 0.01 |
| Fresh ICSI | mothers_age | 34.83 | 4.32 | 30.73 | 5.40 | 0.84 | 32.12 | 4.48 | 30.77 | 5.42 | 0.27 |
|  | smoke_1 | 0.96 | 0.19 | 0.86 | 0.35 | 0.37 | 0.88 | 0.32 | 0.86 | 0.35 | 0.08 |
|  | smoke_2 | 0.02 | 0.14 | 0.11 | 0.32 | 0.37 | 0.09 | 0.28 | 0.11 | 0.31 | 0.08 |
|  | smoke_3 | 0.02 | 0.13 | 0.03 | 0.17 | 0.10 | 0.03 | 0.17 | 0.03 | 0.17 | 0.02 |
|  | prev_preg_1 | 0.66 | 0.47 | 0.38 | 0.49 | 0.57 | 0.37 | 0.48 | 0.39 | 0.49 | 0.03 |
|  | prev_preg_2 | 0.34 | 0.47 | 0.62 | 0.49 | 0.57 | 0.63 | 0.48 | 0.61 | 0.49 | 0.03 |
|  | remoteness_1 | 0.82 | 0.39 | 0.73 | 0.45 | 0.22 | 0.75 | 0.43 | 0.73 | 0.45 | 0.05 |
|  | remoteness_2 | 0.13 | 0.33 | 0.16 | 0.37 | 0.11 | 0.15 | 0.36 | 0.16 | 0.37 | 0.03 |
|  | remoteness_3 | 0.04 | 0.21 | 0.08 | 0.26 | 0.13 | 0.07 | 0.25 | 0.08 | 0.26 | 0.03 |
|  | remoteness_4 | 0.01 | 0.10 | 0.02 | 0.13 | 0.07 | 0.02 | 0.12 | 0.02 | 0.13 | 0.02 |
|  | remoteness_5 | 0.00 | 0.06 | 0.02 | 0.12 | 0.12 | 0.01 | 0.11 | 0.02 | 0.12 | 0.03 |
|  | IRSD_1 | 0.05 | 0.22 | 0.10 | 0.30 | 0.18 | 0.09 | 0.29 | 0.10 | 0.30 | 0.02 |
|  | IRSD_2 | 0.06 | 0.23 | 0.09 | 0.28 | 0.11 | 0.08 | 0.27 | 0.09 | 0.28 | 0.02 |
|  | IRSD_3 | 0.05 | 0.23 | 0.07 | 0.26 | 0.08 | 0.07 | 0.26 | 0.07 | 0.26 | 0.01 |
|  | IRSD_4 | 0.07 | 0.26 | 0.10 | 0.30 | 0.09 | 0.09 | 0.28 | 0.10 | 0.30 | 0.04 |
|  | IRSD_5 | 0.09 | 0.29 | 0.10 | 0.30 | 0.04 | 0.10 | 0.30 | 0.10 | 0.30 | 0.01 |
|  | IRSD_6 | 0.10 | 0.30 | 0.11 | 0.31 | 0.02 | 0.11 | 0.31 | 0.11 | 0.31 | 0.00 |
|  | IRSD_7 | 0.10 | 0.29 | 0.09 | 0.28 | 0.03 | 0.10 | 0.29 | 0.09 | 0.28 | 0.03 |
|  | IRSD_8 | 0.13 | 0.34 | 0.11 | 0.31 | 0.07 | 0.12 | 0.32 | 0.11 | 0.31 | 0.02 |
|  | IRSD_9 | 0.16 | 0.37 | 0.12 | 0.32 | 0.13 | 0.13 | 0.33 | 0.12 | 0.32 | 0.03 |
|  | IRSD_10 | 0.18 | 0.38 | 0.11 | 0.31 | 0.21 | 0.12 | 0.33 | 0.11 | 0.31 | 0.04 |
|  | IRSD_11 | 0.00 | 0.06 | 0.02 | 0.13 | 0.12 | 0.01 | 0.11 | 0.02 | 0.12 | 0.04 |
|  | parent_cancer_1 | 0.96 | 0.19 | 0.99 | 0.11 | 0.15 | 0.98 | 0.14 | 0.99 | 0.11 | 0.05 |
|  | parent_cancer_2 | 0.04 | 0.19 | 0.01 | 0.11 | 0.15 | 0.02 | 0.14 | 0.01 | 0.11 | 0.05 |
|  | diabetes_history_1 | 0.88 | 0.33 | 0.87 | 0.34 | 0.04 | 0.86 | 0.35 | 0.87 | 0.34 | 0.02 |
|  | diabetes_history_2 | 0.02 | 0.15 | 0.03 | 0.18 | 0.07 | 0.03 | 0.18 | 0.03 | 0.18 | 0.01 |
|  | diabetes_history_3 | 0.10 | 0.30 | 0.10 | 0.30 | 0.00 | 0.11 | 0.31 | 0.10 | 0.30 | 0.02 |
| Frozen IVF | mothers_age | 36.11 | 4.54 | 30.73 | 5.40 | 1.08 | 32.31 | 4.27 | 30.76 | 5.42 | 0.32 |
|  | smoke_1 | 0.97 | 0.18 | 0.86 | 0.35 | 0.40 | 0.87 | 0.34 | 0.86 | 0.35 | 0.04 |
|  | smoke_2 | 0.02 | 0.13 | 0.11 | 0.32 | 0.40 | 0.10 | 0.30 | 0.11 | 0.32 | 0.04 |
|  | smoke_3 | 0.02 | 0.12 | 0.03 | 0.17 | 0.10 | 0.03 | 0.18 | 0.03 | 0.17 | 0.00 |
|  | prev_preg_1 | 0.45 | 0.50 | 0.38 | 0.49 | 0.13 | 0.36 | 0.48 | 0.38 | 0.49 | 0.04 |
|  | prev_preg_2 | 0.55 | 0.50 | 0.62 | 0.49 | 0.13 | 0.64 | 0.48 | 0.62 | 0.49 | 0.04 |
|  | remoteness_1 | 0.82 | 0.38 | 0.73 | 0.45 | 0.24 | 0.74 | 0.44 | 0.73 | 0.45 | 0.02 |
|  | remoteness_2 | 0.12 | 0.32 | 0.16 | 0.37 | 0.14 | 0.16 | 0.37 | 0.16 | 0.37 | 0.01 |
|  | remoteness_3 | 0.04 | 0.21 | 0.08 | 0.26 | 0.13 | 0.08 | 0.27 | 0.08 | 0.26 | 0.00 |
|  | remoteness_4 | 0.01 | 0.10 | 0.02 | 0.13 | 0.07 | 0.02 | 0.13 | 0.02 | 0.13 | 0.01 |
|  | remoteness_5 | 0.00 | 0.07 | 0.02 | 0.12 | 0.11 | 0.01 | 0.10 | 0.02 | 0.12 | 0.06 |
|  | IRSD_1 | 0.05 | 0.21 | 0.10 | 0.30 | 0.20 | 0.10 | 0.30 | 0.10 | 0.30 | 0.01 |
|  | IRSD_2 | 0.05 | 0.22 | 0.09 | 0.28 | 0.14 | 0.09 | 0.29 | 0.09 | 0.28 | 0.03 |
|  | IRSD_3 | 0.04 | 0.21 | 0.07 | 0.26 | 0.12 | 0.06 | 0.24 | 0.07 | 0.26 | 0.04 |
|  | IRSD_4 | 0.07 | 0.26 | 0.10 | 0.30 | 0.09 | 0.09 | 0.29 | 0.10 | 0.30 | 0.02 |
|  | IRSD_5 | 0.08 | 0.27 | 0.10 | 0.30 | 0.08 | 0.09 | 0.29 | 0.10 | 0.30 | 0.03 |
|  | IRSD_6 | 0.10 | 0.30 | 0.11 | 0.31 | 0.02 | 0.10 | 0.31 | 0.11 | 0.31 | 0.01 |
|  | IRSD_7 | 0.10 | 0.30 | 0.09 | 0.28 | 0.03 | 0.09 | 0.28 | 0.09 | 0.28 | 0.00 |
|  | IRSD_8 | 0.13 | 0.34 | 0.11 | 0.31 | 0.06 | 0.11 | 0.32 | 0.11 | 0.31 | 0.01 |
|  | IRSD_9 | 0.16 | 0.37 | 0.12 | 0.32 | 0.13 | 0.12 | 0.33 | 0.12 | 0.32 | 0.02 |
|  | IRSD_10 | 0.21 | 0.41 | 0.11 | 0.31 | 0.28 | 0.12 | 0.32 | 0.11 | 0.31 | 0.03 |
|  | IRSD_11 | 0.00 | 0.07 | 0.02 | 0.13 | 0.11 | 0.01 | 0.10 | 0.02 | 0.13 | 0.06 |
|  | parent_cancer_1 | 0.97 | 0.17 | 0.99 | 0.11 | 0.11 | 0.99 | 0.12 | 0.99 | 0.11 | 0.02 |
|  | parent_cancer_2 | 0.03 | 0.17 | 0.01 | 0.11 | 0.11 | 0.01 | 0.12 | 0.01 | 0.11 | 0.02 |
|  | diabetes_history_1 | 0.76 | 0.43 | 0.87 | 0.34 | 0.27 | 0.86 | 0.35 | 0.86 | 0.34 | 0.01 |
|  | diabetes_history_2 | 0.03 | 0.18 | 0.03 | 0.18 | 0.00 | 0.03 | 0.18 | 0.03 | 0.18 | 0.01 |
|  | diabetes_history_3 | 0.21 | 0.40 | 0.10 | 0.30 | 0.29 | 0.10 | 0.31 | 0.10 | 0.30 | 0.01 |
| Frozen ICSI | mothers_age | 35.66 | 4.38 | 30.73 | 5.40 | 1.00 | 32.17 | 4.35 | 30.76 | 5.42 | 0.29 |
|  | smoke_1 | 0.98 | 0.15 | 0.86 | 0.35 | 0.45 | 0.88 | 0.33 | 0.86 | 0.35 | 0.06 |
|  | smoke_2 | 0.02 | 0.12 | 0.11 | 0.32 | 0.41 | 0.09 | 0.28 | 0.11 | 0.32 | 0.08 |
|  | smoke_3 | 0.01 | 0.09 | 0.03 | 0.17 | 0.17 | 0.04 | 0.18 | 0.03 | 0.17 | 0.02 |
|  | prev_preg_1 | 0.50 | 0.50 | 0.38 | 0.49 | 0.24 | 0.36 | 0.48 | 0.39 | 0.49 | 0.04 |
|  | prev_preg_2 | 0.50 | 0.50 | 0.62 | 0.49 | 0.24 | 0.64 | 0.48 | 0.61 | 0.49 | 0.04 |
|  | remoteness_1 | 0.83 | 0.38 | 0.73 | 0.45 | 0.25 | 0.74 | 0.44 | 0.73 | 0.45 | 0.02 |
|  | remoteness_2 | 0.12 | 0.32 | 0.16 | 0.37 | 0.13 | 0.16 | 0.37 | 0.16 | 0.37 | 0.01 |
|  | remoteness_3 | 0.04 | 0.20 | 0.08 | 0.26 | 0.15 | 0.07 | 0.26 | 0.08 | 0.26 | 0.01 |
|  | remoteness_4 | 0.01 | 0.09 | 0.02 | 0.13 | 0.09 | 0.02 | 0.13 | 0.02 | 0.13 | 0.00 |
|  | remoteness_5 | 0.00 | 0.06 | 0.02 | 0.12 | 0.12 | 0.01 | 0.11 | 0.02 | 0.12 | 0.03 |
|  | IRSD_1 | 0.05 | 0.21 | 0.10 | 0.30 | 0.19 | 0.10 | 0.30 | 0.10 | 0.30 | 0.00 |
|  | IRSD_2 | 0.05 | 0.22 | 0.09 | 0.28 | 0.13 | 0.08 | 0.28 | 0.09 | 0.28 | 0.00 |
|  | IRSD_3 | 0.05 | 0.22 | 0.07 | 0.26 | 0.10 | 0.07 | 0.26 | 0.07 | 0.26 | 0.00 |
|  | IRSD_4 | 0.07 | 0.25 | 0.10 | 0.30 | 0.11 | 0.09 | 0.29 | 0.10 | 0.30 | 0.03 |
|  | IRSD_5 | 0.08 | 0.28 | 0.10 | 0.30 | 0.06 | 0.10 | 0.30 | 0.10 | 0.30 | 0.01 |
|  | IRSD_6 | 0.10 | 0.30 | 0.11 | 0.31 | 0.02 | 0.11 | 0.31 | 0.11 | 0.31 | 0.01 |
|  | IRSD_7 | 0.10 | 0.29 | 0.09 | 0.28 | 0.03 | 0.09 | 0.28 | 0.09 | 0.28 | 0.01 |
|  | IRSD_8 | 0.13 | 0.34 | 0.11 | 0.31 | 0.07 | 0.11 | 0.32 | 0.11 | 0.31 | 0.01 |
|  | IRSD_9 | 0.17 | 0.38 | 0.12 | 0.32 | 0.17 | 0.12 | 0.32 | 0.12 | 0.32 | 0.01 |
|  | IRSD_10 | 0.19 | 0.39 | 0.11 | 0.31 | 0.24 | 0.11 | 0.32 | 0.11 | 0.31 | 0.02 |
|  | IRSD_11 | 0.00 | 0.06 | 0.02 | 0.13 | 0.12 | 0.01 | 0.11 | 0.02 | 0.12 | 0.03 |
|  | parent_cancer_1 | 0.96 | 0.20 | 0.99 | 0.11 | 0.18 | 0.98 | 0.13 | 0.99 | 0.11 | 0.03 |
|  | parent_cancer_2 | 0.04 | 0.20 | 0.01 | 0.11 | 0.18 | 0.02 | 0.13 | 0.01 | 0.11 | 0.03 |
|  | diabetes_history_1 | 0.79 | 0.40 | 0.87 | 0.34 | 0.19 | 0.86 | 0.35 | 0.86 | 0.34 | 0.02 |
|  | diabetes_history_2 | 0.03 | 0.18 | 0.03 | 0.18 | 0.00 | 0.04 | 0.19 | 0.03 | 0.18 | 0.02 |
|  | diabetes_history_3 | 0.17 | 0.38 | 0.10 | 0.30 | 0.21 | 0.11 | 0.31 | 0.10 | 0.30 | 0.01 |
| SD: Standard deviation; MAR: Medically Assisted Reproduction; IVF: In-Vitro Fertilisation; ICSI: Intracytoplasmic Sperm Injection; OI/IUI: Ovulation Induction/Intrauterine Insemination.  These map onto confounding variables as follows:  Mothers_age: Maternal age at birth (numeric); smoke: History of smoking (1 = No, 2 = Yes, 3 = Missing); prev_preg: Any history of previous pregnancy (1 = Yes, 2 = No); remoteness: Remoteness of residence (1-4 in order of increasing remoteness, 5 = Missing); IRSD: Index of Relative Socioeconomic Disadvantage of Area of Residence Decile (1-10 in order of decreasing disadvantage, 11 = Missing); parent_cancer: History of cancer in either parent (1 = No, 2 = Yes); diabetes_history: Maternal history of diabetes (1 = No, 2 = Yes, 3 = Missing). | | | | | | | | | | | |

#### Supplementary Table 4: Cohort demographics table by treatment type for the primary cohort (IVF, ICSI, Fresh and frozen embryo transfers)

|  | **IVF** | **ICSI** | **Frozen ART** | **Fresh ART** | **Frozen IVF** | **Frozen ICSI** | **Fresh IVF** | **Fresh ICSI** |
| --- | --- | --- | --- | --- | --- | --- | --- | --- |
| **N** | 40,463 | 53,344 | 41,344 | 67,388 | 17,274 | 20,914 | 23,189 | 32,430 |
| **Child sex** |  |  |  |  |  |  |  |  |
| Male | 21,455 (53.0%) | 26,748 (50.1%) | 20,948 (50.7%) | 34,956 (51.9%) | 8,922 (51.6%) | 10,383 (49.6%) | 12,533 (54.0%) | 16,365 (50.5%) |
| Female | 18,785 (46.4%) | 26,288 (49.3%) | 20,158 (48.8%) | 32,007 (47.5%) | 8,254 (47.8%) | 10,425 (49.8%) | 10,531 (45.4%) | 15,863 (48.9%) |
| Missing | 223 (0.6%) | 308 (0.6%) | 238 (0.6%) | 425 (0.6%) | 98 (0.6%) | 106 (0.5%) | 125 (0.5%) | 202 (0.6%) |
| **Year of birth** |  |  |  |  |  |  |  |  |
| 1991-1994 | NA | NA | 42 (0.1%) | 211 (0.3%) | NA | NA | NA | NA |
| 1995-1999 | NA | NA | 145 (0.4%) | 1,104 (1.6%) | NA | NA | NA | NA |
| 2000-2004 | NA | NA | 1,749 (4.2%) | 6,174 (9.2%) | NA | NA | NA | NA |
| 2005-2009 | 8,278 (20.5%) | 6,535 (12.3%) | 4,840 (11.7%) | 15,473 (23.0%) | 2,496 (14.4%) | 1,124 (5.4%) | 5,782 (24.9%) | 5,411 (16.7%) |
| 2010-2014 | 15,194 (37.6%) | 22,015 (41.3%) | 12,804 (31.0%) | 24,405 (36.2%) | 5,751 (33.3%) | 7,053 (33.7%) | 9,443 (40.7%) | 14,962 (46.1%) |
| 2015-2019 | 16,991 (42.0%) | 24,794 (46.5%) | 21,764 (52.6%) | 20,021 (29.7%) | 9,027 (52.3%) | 12,737 (60.9%) | 7,964 (34.3%) | 12,057 (37.2%) |
| **State of residence** |  |  |  |  |  |  |  |  |
| Australian Capital Territory | 352 (0.9%) | 433 (0.8%) | 283 (0.7%) | 613 (0.9%) | 125 (0.7%) | 144 (0.7%) | 227 (1.0%) | 289 (0.9%) |
| New South Wales | 18,054 (44.6%) | 19,092 (35.8%) | 16,613 (40.2%) | 28,762 (42.7%) | 7,282 (42.2%) | 7,225 (34.5%) | 10,772 (46.5%) | 11,867 (36.6%) |
| Queensland | 6,582 (16.3%) | 8,080 (15.1%) | 6,023 (14.6%) | 8,639 (12.8%) | 2,978 (17.2%) | 3,045 (14.6%) | 3,604 (15.5%) | 5,035 (15.5%) |
| South Australia | 1,972 (4.9%) | 4,684 (8.8%) | 2,501 (6.0%) | 6,449 (9.6%) | 738 (4.3%) | 1,460 (7.0%) | 1,234 (5.3%) | 3,224 (9.9%) |
| Tasmania | 845 (2.1%) | 932 (1.7%) | 973 (2.4%) | 1,010 (1.5%) | 454 (2.6%) | 441 (2.1%) | 391 (1.7%) | 491 (1.5%) |
| Victoria | 7,867 (19.4%) | 15,926 (29.9%) | 10,662 (25.8%) | 15,846 (23.5%) | 3,508 (20.3%) | 6,705 (32.1%) | 4,359 (18.8%) | 9,221 (28.4%) |
| Western Australia | 4,791 (11.8%) | 4,197 (7.9%) | 4,289 (10.4%) | 6,069 (9.0%) | 2,189 (12.7%) | 1,894 (9.1%) | 2,602 (11.2%) | 2,303 (7.1%) |
| **Residential remoteness** |  |  |  |  |  |  |  |  |
| Major cities of Australia | 33,430 (82.6%) | 43,808 (82.1%) | 33,981 (82.2%) | 54,848 (81.4%) | 14,222 (82.3%) | 17,330 (82.9%) | 19,208 (82.8%) | 26,478 (81.6%) |
| Inner regional Australia | 4,595 (11.4%) | 6,569 (12.3%) | 4,966 (12.0%) | 7,862 (11.7%) | 2,037 (11.8%) | 2,509 (12.0%) | 2,558 (11.0%) | 4,060 (12.5%) |
| Outer regional Australia | 1,862 (4.6%) | 2,293 (4.3%) | 1,710 (4.1%) | 2,996 (4.4%) | 772 (4.5%) | 834 (4.0%) | 1,090 (4.7%) | 1,459 (4.5%) |
| Remote Australia | 275 (0.7%) | 345 (0.6%) | 248 (0.6%) | 456 (0.7%) | 124 (0.7%) | 114 (0.5%) | 151 (0.7%) | 231 (0.7%) |
| Very remote Australia | 109 (0.3%) | 129 (0.2%) | 91 (0.2%) | 182 (0.3%) | * | * | * | * |
| Missing | 192 (0.5%) | 200 (0.4%) | 348 (0.8%) | 1,044 (1.5%) | * | * | * | * |
| **Index of Relative Socioeconomic Disadvantage quintile** | | | | |  |  |  |  |
| 1 (most disadvantaged) | 4,133 (10.2%) | 5,557 (10.4%) | 4,100 (9.9%) | 7,260 (10.8%) | 1,673 (9.7%) | 2,073 (9.9%) | 2,460 (10.6%) | 3,484 (10.7%) |
| 2 | 4,894 (12.1%) | 6,619 (12.4%) | 4,884 (11.8%) | 8,525 (12.7%) | 2,046 (11.8%) | 2,469 (11.8%) | 2,848 (12.3%) | 4,150 (12.8%) |
| 3 | 7,532 (18.6%) | 10,093 (18.9%) | 7,519 (18.2%) | 12,334 (18.3%) | 3,143 (18.2%) | 3,895 (18.6%) | 4,389 (18.9%) | 6,198 (19.1%) |
| 4 | 9,062 (22.4%) | 12,232 (22.9%) | 9,235 (22.3%) | 14,680 (21.8%) | 3,927 (22.7%) | 4,785 (22.9%) | 5,135 (22.1%) | 7,447 (23.0%) |
| 5 (least disadvantaged) | 14,644 (36.2%) | 18,630 (34.9%) | 15,246 (36.9%) | 23,534 (34.9%) | 6,400 (37.0%) | 7,606 (36.4%) | 8,244 (35.6%) | 11,024 (34.0%) |
| Missing | 198 (0.5%) | 213 (0.4%) | 360 (0.9%) | 1,055 (1.6%) | 85 (0.5%) | 86 (0.4%) | 113 (0.5%) | 127 (0.4%) |
| **Gestational age (weeks)** |  |  |  |  |  |  |  |  |
| Mean (SD) | 38.4 (1.9) | 38.4 (1.9) | 38.5 (1.8) | 38.5 (2.0) | 38.4 (1.8) | 38.4 (1.8) | 38.4 (2.0) | 38.5 (1.9) |
| Median [IQR] | 39.0 [38.0-40.0] | 39.0 [38.0-40.0] | 39.0 [38.0-40.0] | 39.0 [38.0-40.0] | 39.0 [38.0-39.0] | 39.0 [38.0-39.0] | 39.0 [38.0-40.0] | 39.0 [38.0-40.0] |
| **Birthweight (grams)** |  |  |  |  |  |  |  |  |
| Mean (SD) | 3,308.1 (575.3) | 3,322.3 (569.0) | 3,381.5 (564.2) | 3,271.9 (575.9) | 3,369.2 (568.9) | 3,384.9 (558.3) | 3,262.5 (575.9) | 3,282.0 (572.1) |
| Median [IQR] | 3,340 [3,010-3,660] | 3,350 [3,020-3,670] | 3,400 [3,080- 3,730] | 3,300 [2,975-3,625] | 3,390 [3,070- 3,715] | 3,405 [3,090- 3,730] | 3,300 [2,970- 3,612] | 3,310 [2,984- 3,630] |
| **Maternal age at child's birth** | |  |  |  |  |  |  |  |
| Mean (SD) | 35.4 (4.4) | 35.2 (4.4) | 35.8 (4.5) | 34.8 (4.3) | 36.1 (4.5) | 35.7 (4.4) | 34.9 (4.2) | 34.8 (4.3) |
| Median [IQR] | 35.0 [32.0-38.0] | 35.0 [32.0-38.0] | 36.0 [33.0-39.0] | 35.0 [32.0-38.0] | 36.0 [33.0-39.0] | 36.0 [33.0-39.0] | 35.0 [32.0-38.0] | 35.0 [32.0-38.0] |
| **Mother previous pregnancy (> 20 weeks)** | | |  |  |  |  |  |  |
| Yes | 17,974 (44.4%) | 21,516 (40.3%) | 21,408 (51.8%) | 23,057 (34.2%) | 9,500 (55.0%) | 10,433 (49.9%) | 8,474 (36.5%) | 11,083 (34.2%) |
| **Record of maternal smoking** | | |  |  |  |  |  |  |
| No record of smoking | 38,749 (95.8%) | 51,613 (96.8%) | 39,635 (95.9%) | 61,556 (91.3%) | 16,710 (96.7%) | 20,424 (97.7%) | 22,039 (95.0%) | 31,189 (96.2%) |
| Record of smoking | 767 (1.9%) | 1,013 (1.9%) | 706 (1.7%) | 1,591 (2.4%) | 293 (1.7%) | 315 (1.5%) | 474 (2.0%) | 698 (2.2%) |
| Not available | 947 (2.3%) | 718 (1.3%) | 1,003 (2.4%) | 4,241 (6.3%) | 271 (1.6%) | 175 (0.8%) | 676 (2.9%) | 543 (1.7%) |
| **At least one parent with recorded cancer before conception** | | | | | |  |  |  |
| Yes | 977 (2.4%) | 2,028 (3.8%) | 1,438 (3.5%) | 1,945 (2.9%) | 493 (2.9%) | 853 (4.1%) | 484 (2.1%) | 1,175 (3.6%) |
| **History of maternal diabetes** | |  |  |  |  |  |  |  |
| No | 32,693 (80.8%) | 45,034 (84.4%) | 32,822 (79.4%) | 59,613 (88.5%) | 13,139 (76.1%) | 16,592 (79.3%) | 19,554 (84.3%) | 28,442 (87.7%) |
| Yes | 1,046 (2.6%) | 1,411 (2.6%) | 1,333 (3.2%) | 1,341 (2.0%) | 568 (3.3%) | 700 (3.3%) | 478 (2.1%) | 711 (2.2%) |
| Missing | 6,724 (16.6%) | 6,899 (12.9%) | 7,189 (17.4%) | 6,434 (9.5%) | 3,567 (20.6%) | 3,622 (17.3%) | 3,157 (13.6%) | 3,277 (10.1%) |
| **Time to first observed cancer (years)** | | |  |  |  |  |  |  |
| Mean (SD) | 3.6 (2.6) | 3.1 (2.6) | 3.2 (2.9) | 4.4 (3.6) | 3.0 (2.2) | 2.5 (2.3) | 4.1 (2.8) | 3.3 (2.7) |
| Median [IQR] | 3.3 [1.5-5.3] | 2.2 [0.7-4.8] | 2.2 [1.0-4.9] | 3.5 [1.6-5.8] | 2.8 [1.3-4.6] | 1.5 [0.7-3.9] | 3.4 [2.3-5.4] | 3.1 [0.9-5.1] |
| **Total available person time in study (years)** | | |  |  |  |  |  |  |
| Mean (SD) | 6.7 (3.7) | 6.1 (3.3) | 6.0 (4.0) | 8.4 (4.3) | 5.8 (3.5) | 4.9 (2.9) | 7.4 (3.7) | 6.8 (3.3) |
| Median [IQR] | 6.0 [3.0-10.0] | 6.0 [3.0-9.0] | 5.0 [3.0-8.0] | 8.0 [5.0-12.0] | 5.0 [3.0-8.0] | 4.0 [3.0-7.0] | 7.0 [4.0-10.0] | 7.0 [4.0-10.0] |
| **Died in study period** |  |  |  |  |  |  |  |  |
| Yes | 55 (0.1%) | 94 (0.2%) | 73 (0.2%) | 128 (0.2%) | 21 (0.1%) | 40 (0.2%) | 34 (0.1%) | 54 (0.2%) |
| * Censored to prevent possibility of reidentification through small cell size | | | | | | | | |
| IVF: In-Vitro Fertilisation; ICSI: Intracytoplasmic Sperm Injection; SD: Standard Deviation; IQR: Interquartile Range. | | | | | | | | |

### 2. Supplementary Figures

#### Supplementary Figures 1 A-I: Love plots for balance of confounding variables before and after weighting. These map onto confounding variables as follows: Mothers_age: Maternal age at birth (numeric); smoke: History of smoking (1 = No, 2 = Yes, 3 = Missing); prev_preg: Any history of previous pregnancy (1 = Yes, 2 = No); remoteness: Remoteness of residence (1-4 in order of increasing remoteness, 5 = Missing); IRSD: Index of Relative Socioeconomic Disadvantage of Area of Residence Decile (1-10 in order of decreasing disadvantage, 11 = Missing); parent_cancer: History of cancer in either parent (1 = No, 2 = Yes); diabetes_history: Maternal history of diabetes (1 = No, 2 = Yes, 3 = Missing).

A. Medically Assisted Reproduction (MAR)

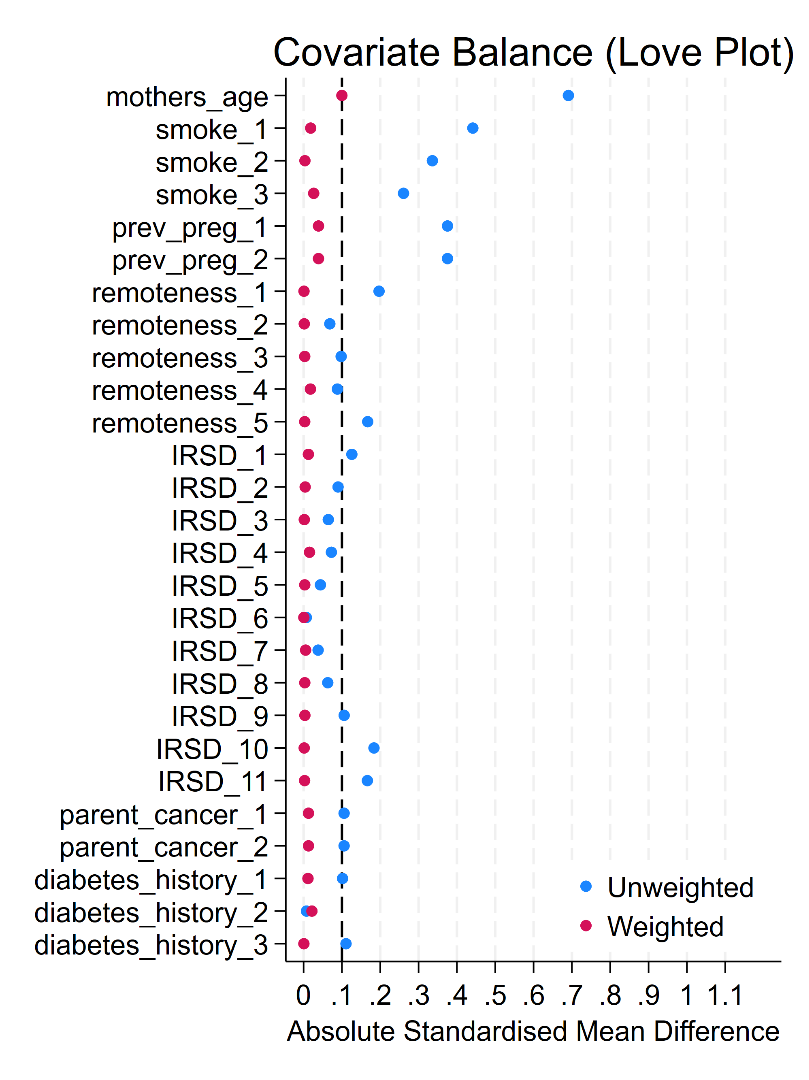

B. Assisted Reproductive Technology (ART)

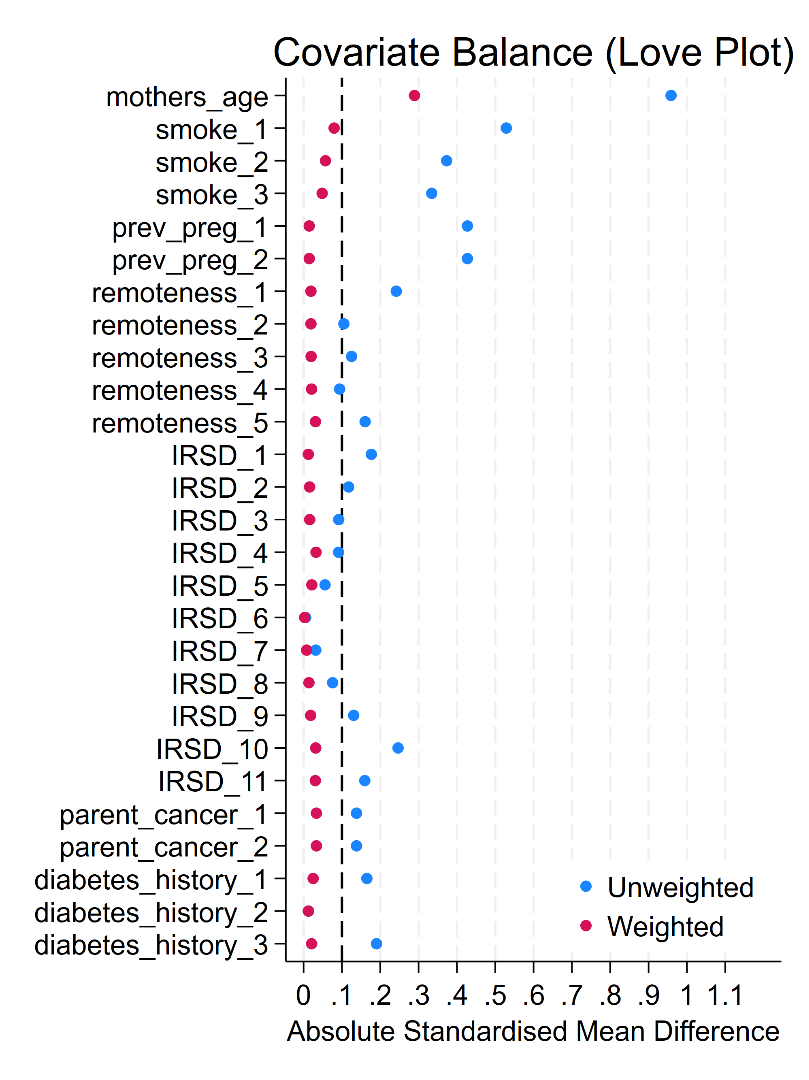

C. Ovulation Induction/Intrauterine Insemination (OI/IUI)

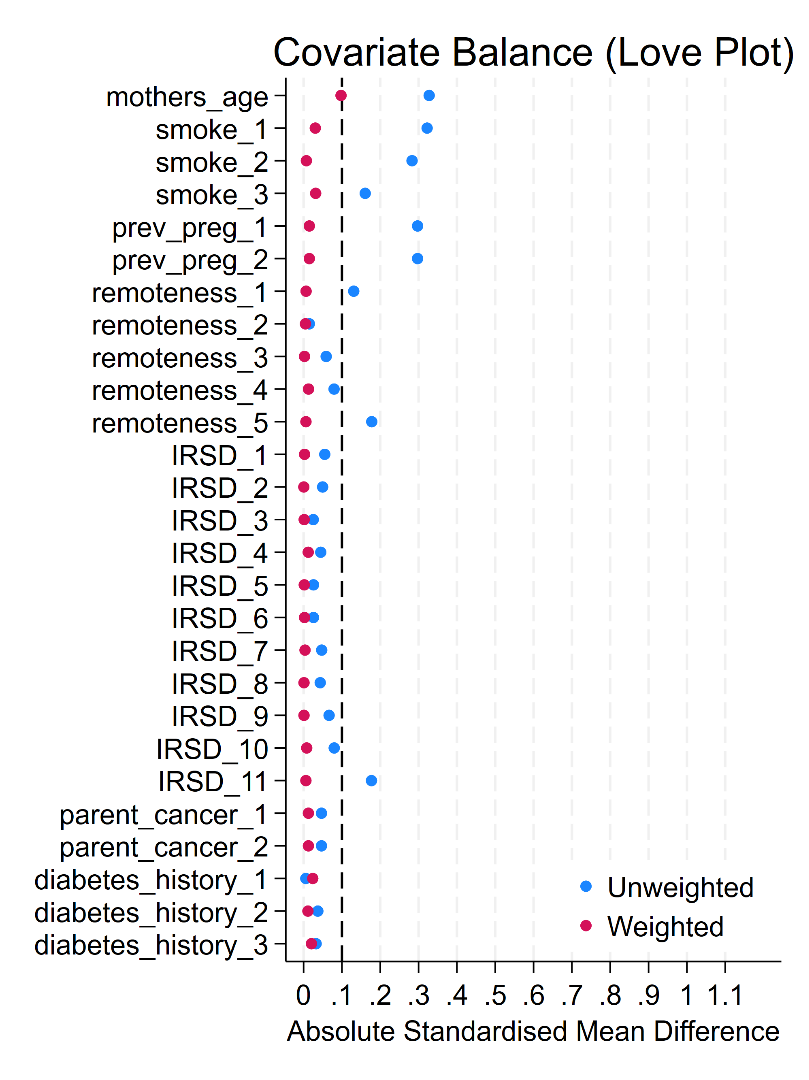

D. In-Vitro Fertilisation (IVF)

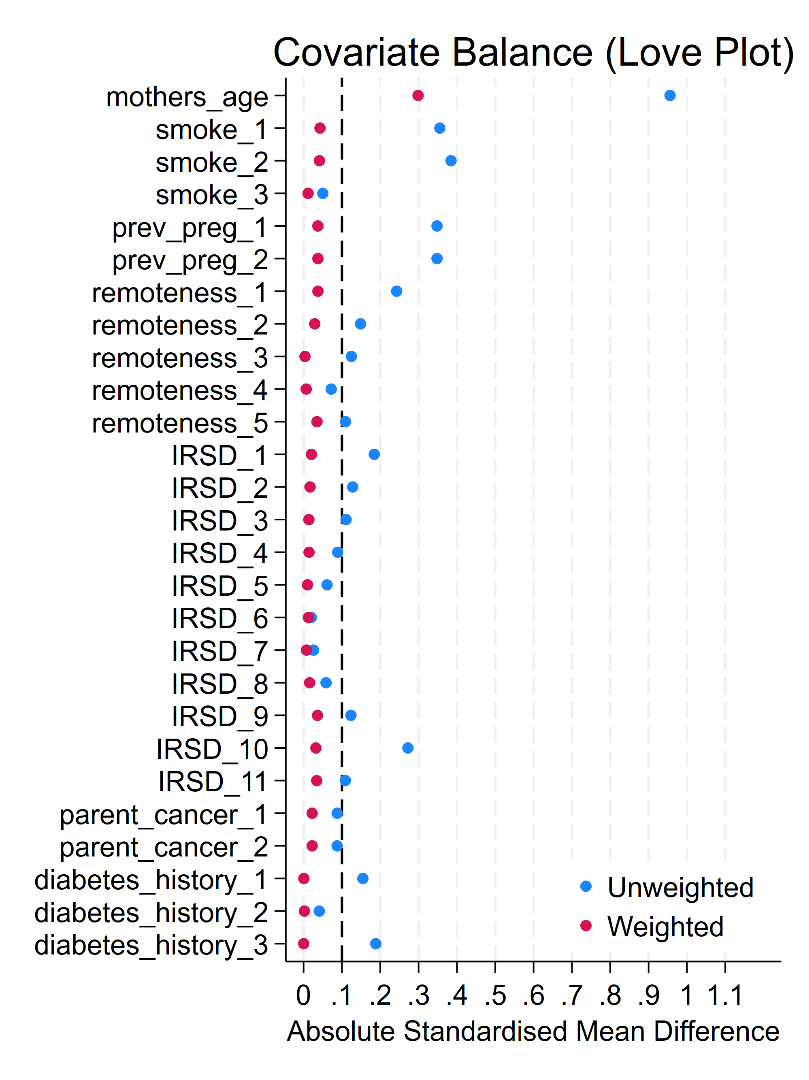

E. Intracytoplasmic Sperm Injection (ICSI)

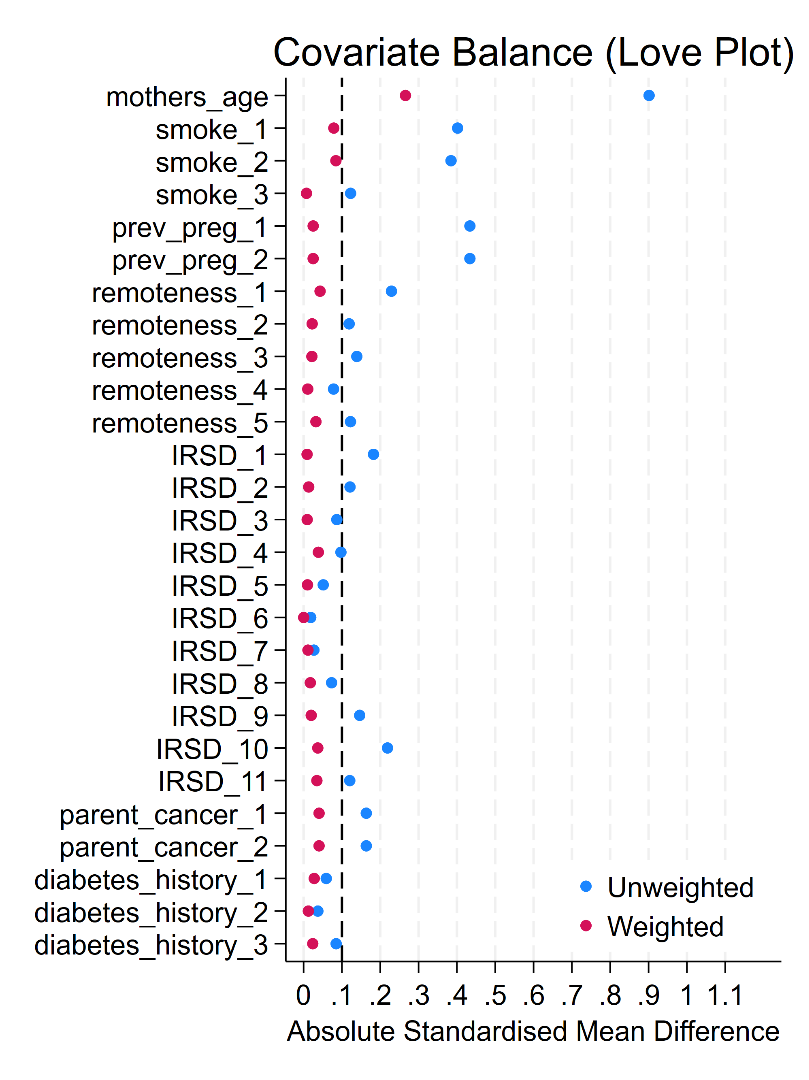

F. Fresh Embryo Transfer

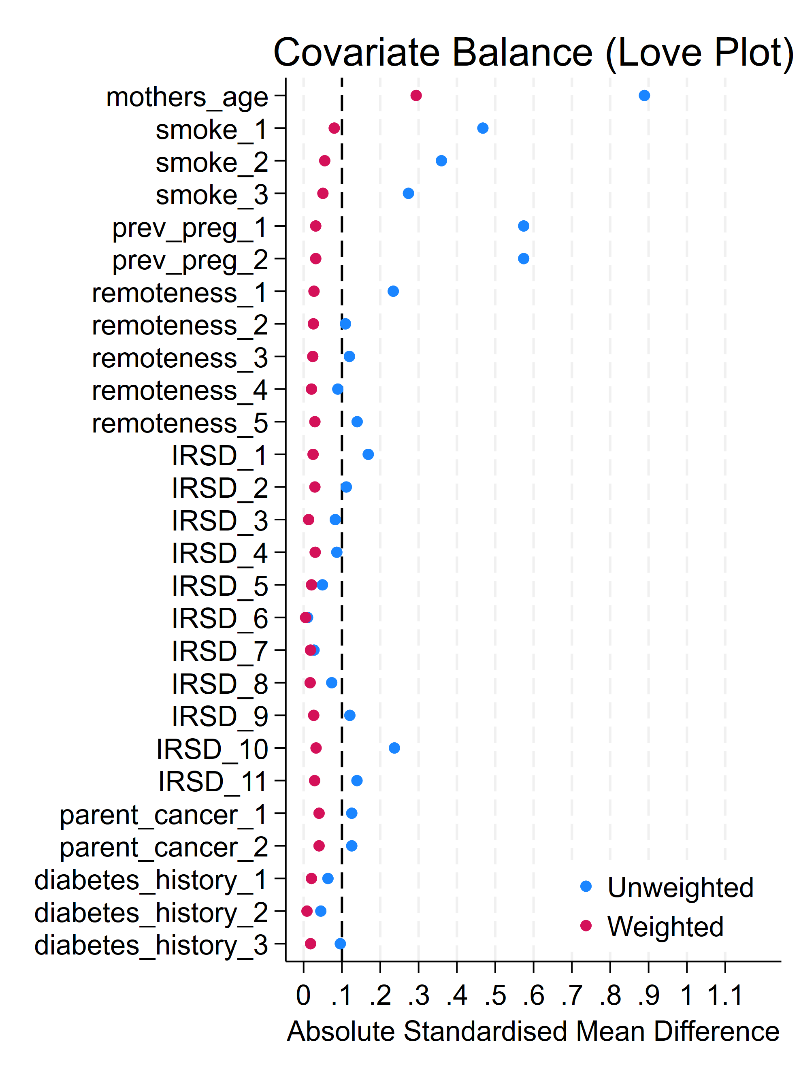

G. Frozen Embryo Transfer

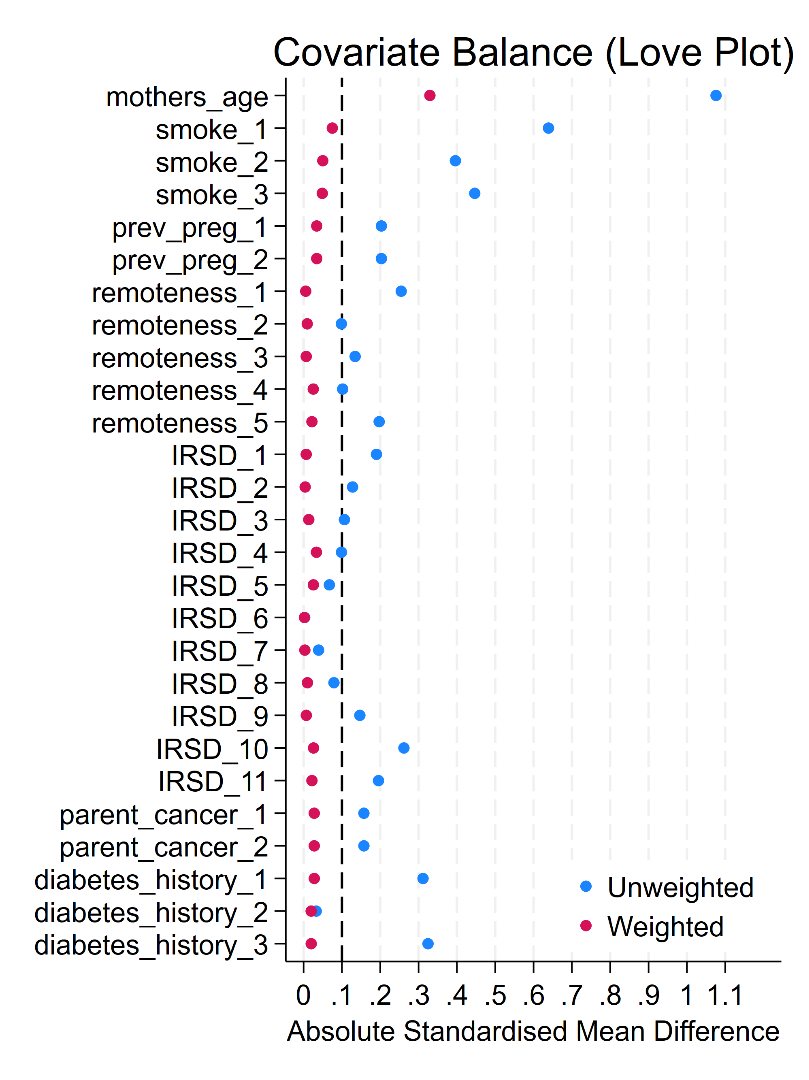

H. Fresh Embryo Transfer In-Vitro Fertilisation (IVF)

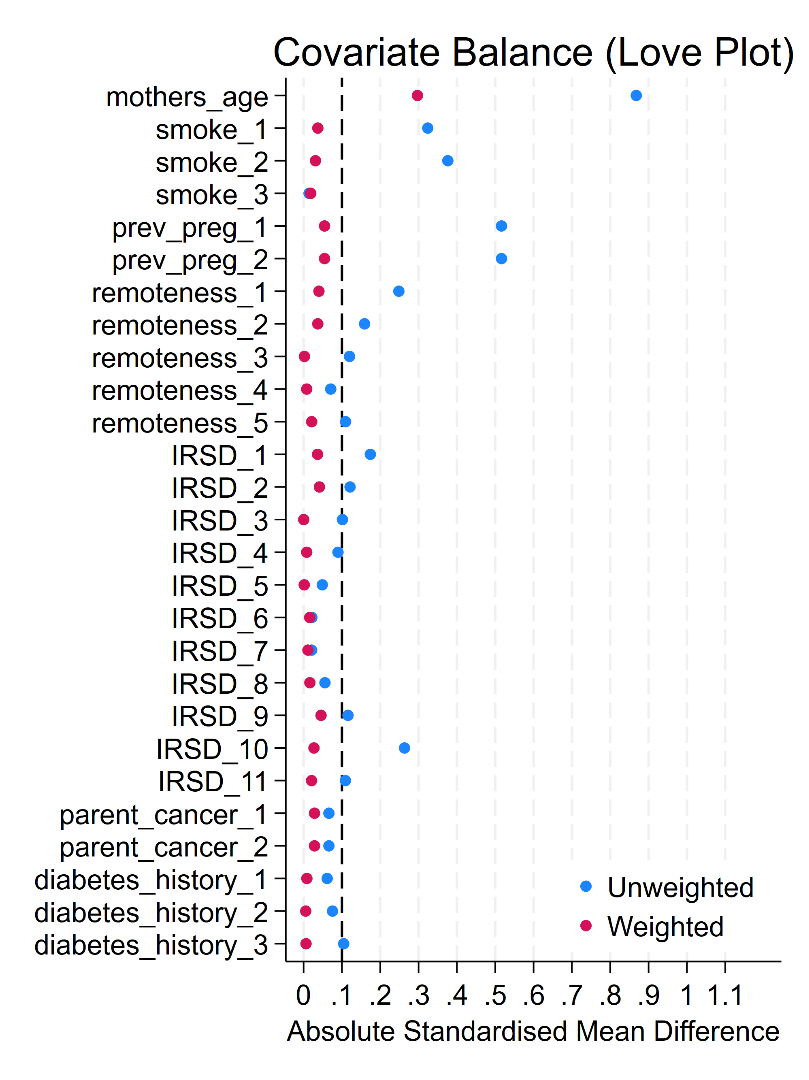

I. Fresh Embryo Transfer Intracytoplasmic Sperm Injection (ICSI)

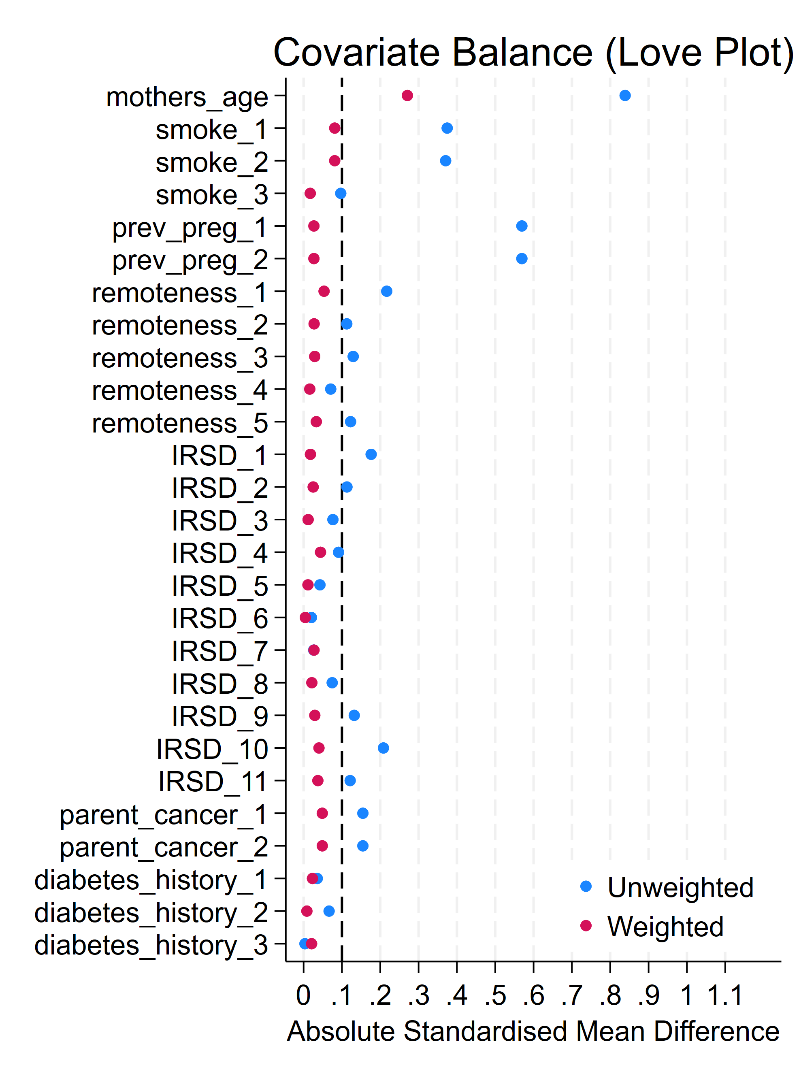

H. Frozen Embryo Transfer In-Vitro Fertilisation (IVF)

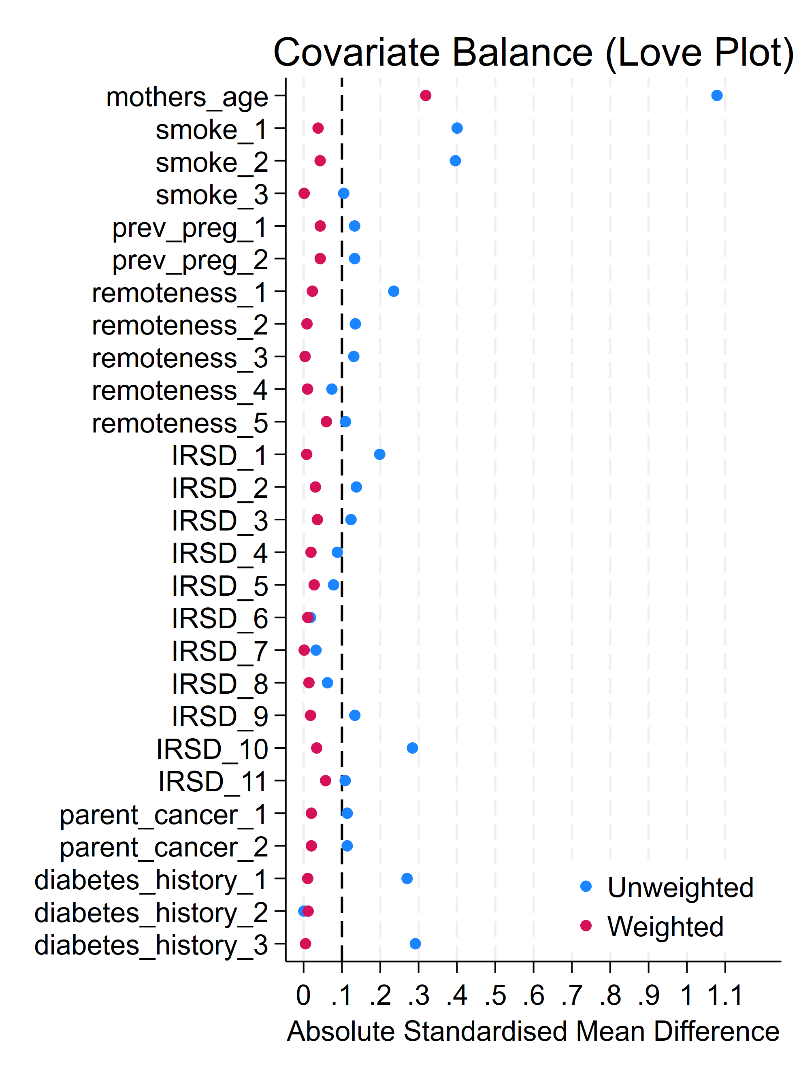

I. Frozen Embryo Transfer Intracytoplasmic Sperm Injection (ICSI)

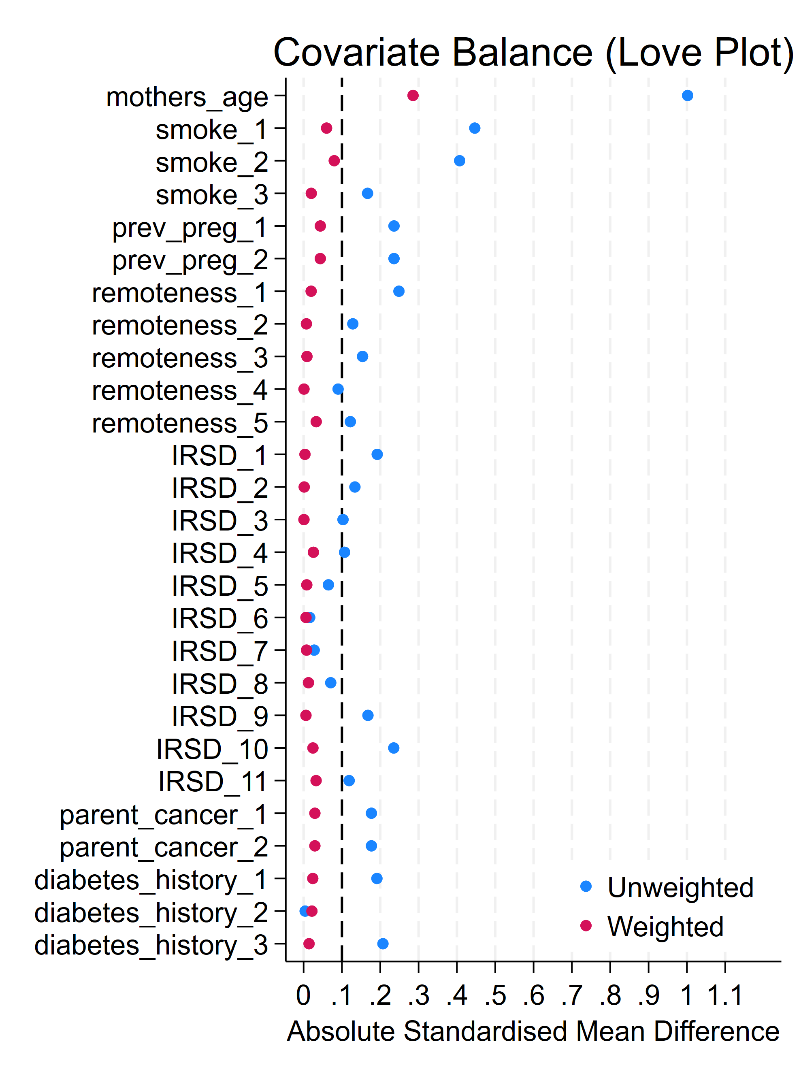

#### Supplementary Figure 2: Standardised incidence ratios (SIRs) splitting assisted reproductive treatments (ART) by in-vitro fertilisation (IVF) and intracytoplasmic sperm injection (ICSI); and by fresh versus frozen embryo transfer.

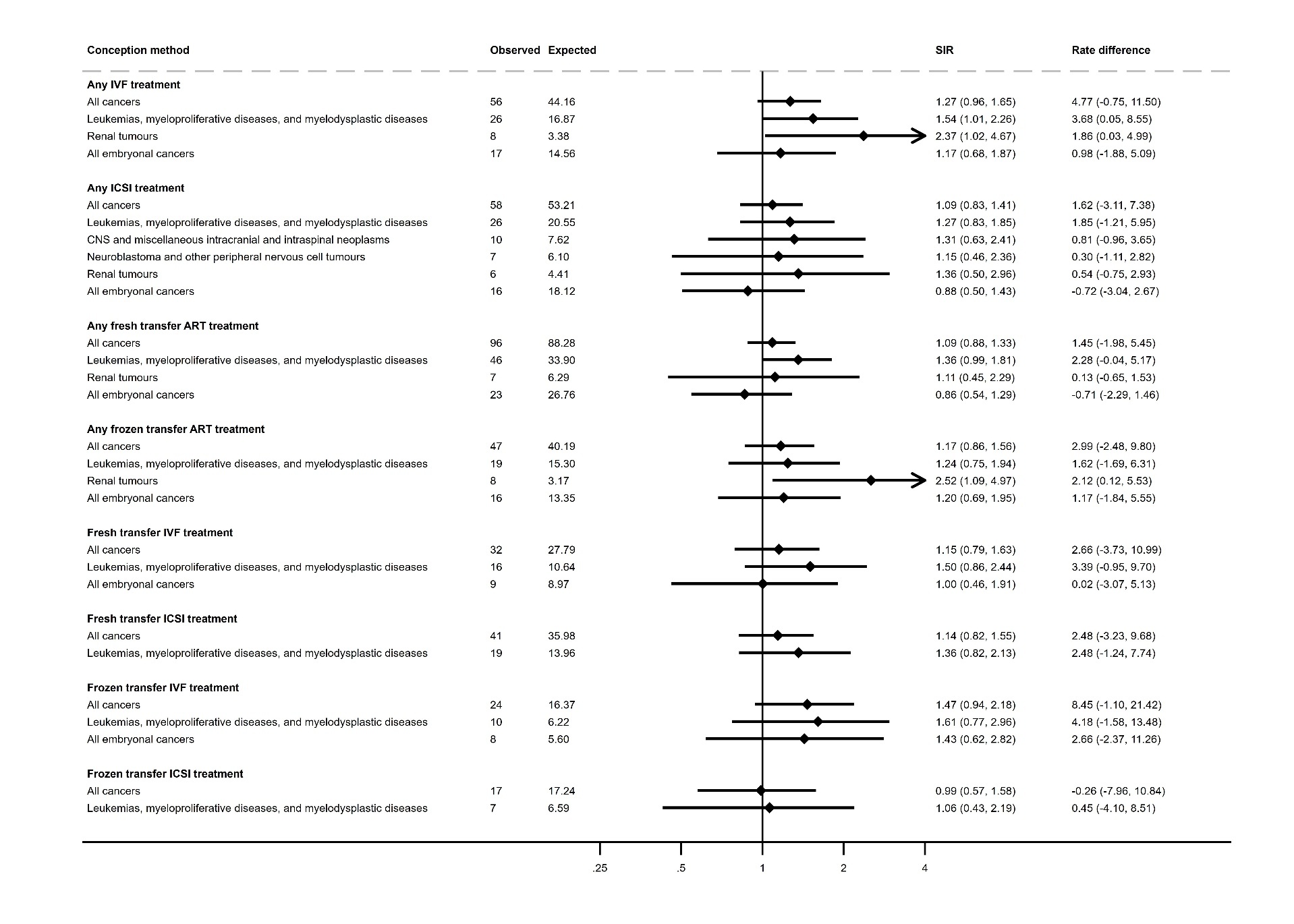

#### Supplementary Figure 3: Standardised incidence ratios (SIRs) for any childhood cancer for children conceived via medically assisted reproduction stratified by age bracket of the child, sex at birth, term or preterm birth, and maternal age.

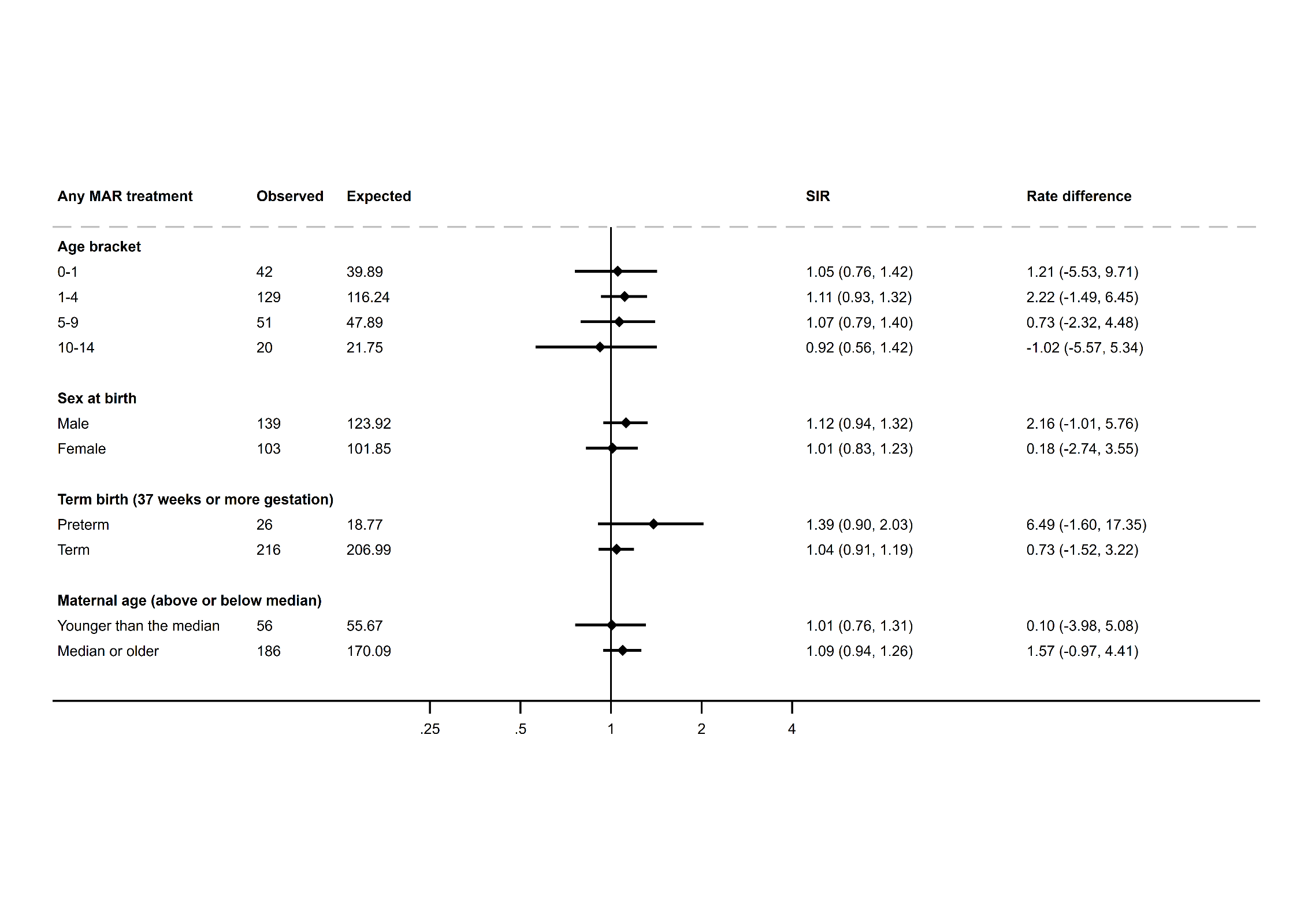

#### Supplementary Figure 4: Standardised incidence ratios (SIRs) for any childhood cancer for children conceived via assisted reproductive technology stratified by age bracket of the child, sex at birth, term or preterm birth, and maternal age.

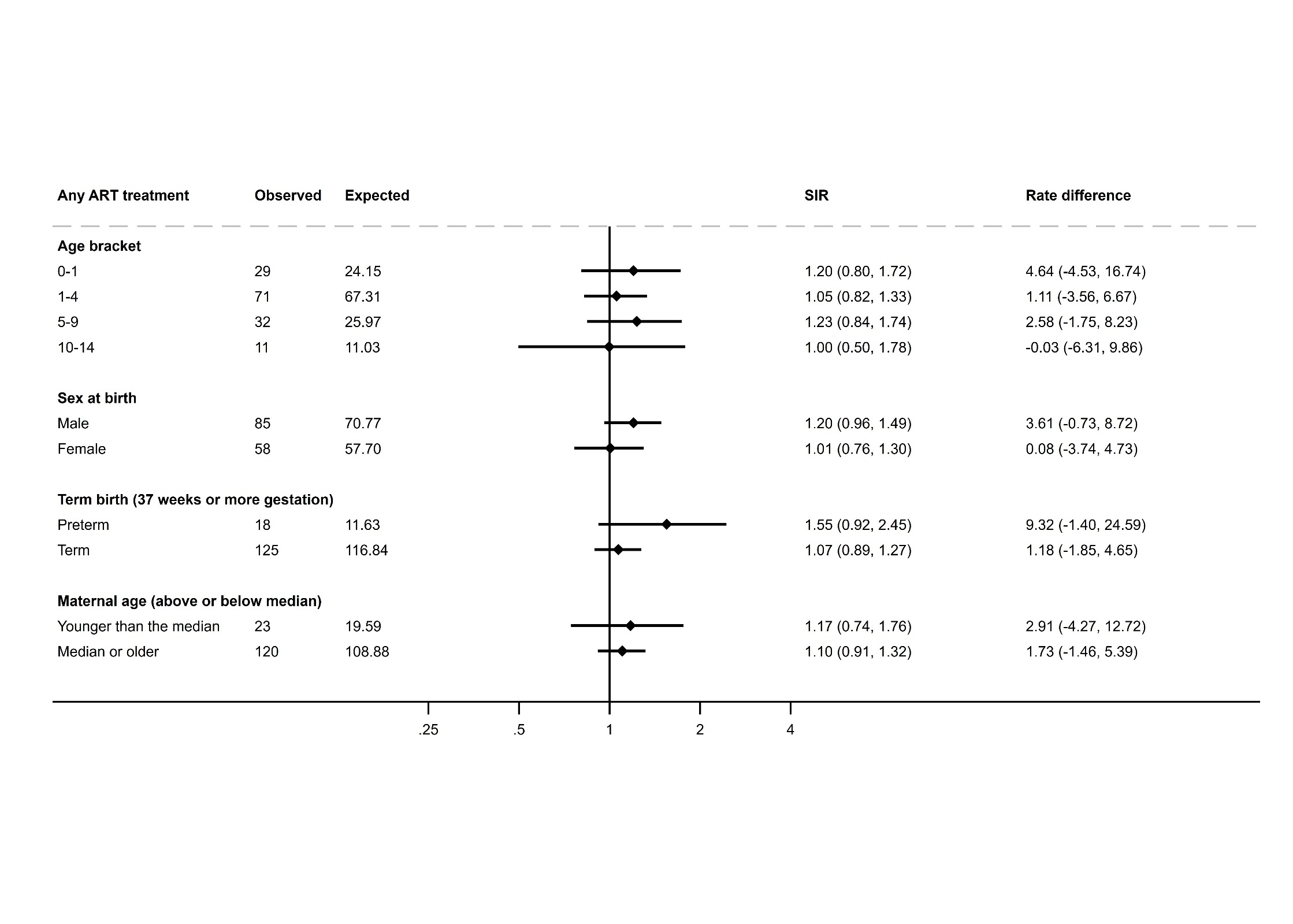

#### Supplementary Figure 5: Standardised incidence ratios (SIRs) for any childhood cancer for children conceived via ovulation induction/intrauterine insemination stratified by age bracket of the child, sex at birth, term or preterm birth, and maternal age.

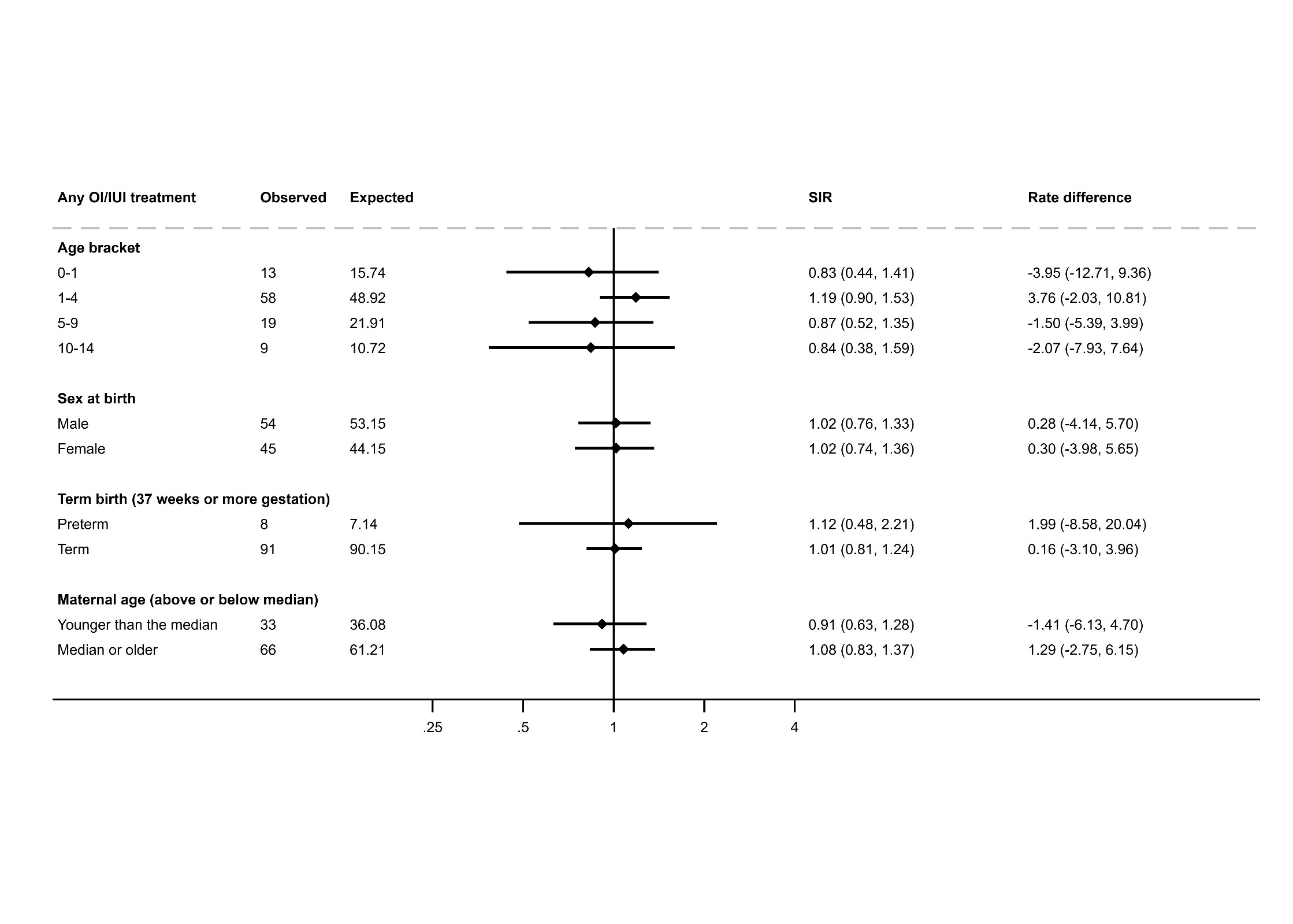

#### Supplementary Figure 6: Hazard ratios splitting assisted reproductive treatments (ART) by in-vitro fertilisation (IVF) and intracytoplasmic sperm injection (ICSI); and by fresh versus frozen embryo transfer.

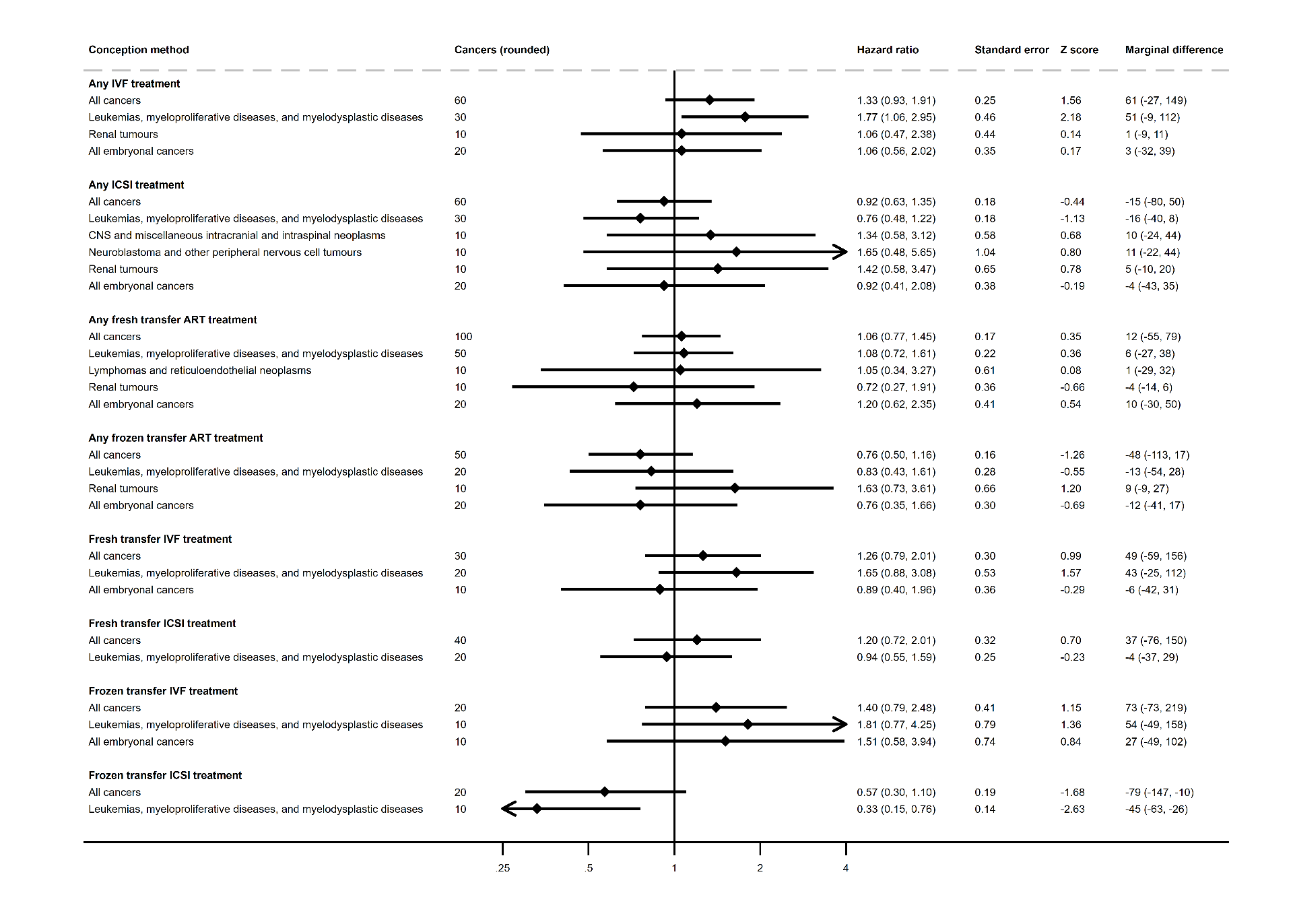

#### Supplementary Figure 7: Hazard ratios and cumulative marginal cancer difference for childhood cancers for children conceived using any medically assisted reproduction (MAR), any assisted reproductive treatment (ART) and any ovulation induction/intrauterine insemination (OI/IUI) - doubly-robust control for maternal age.

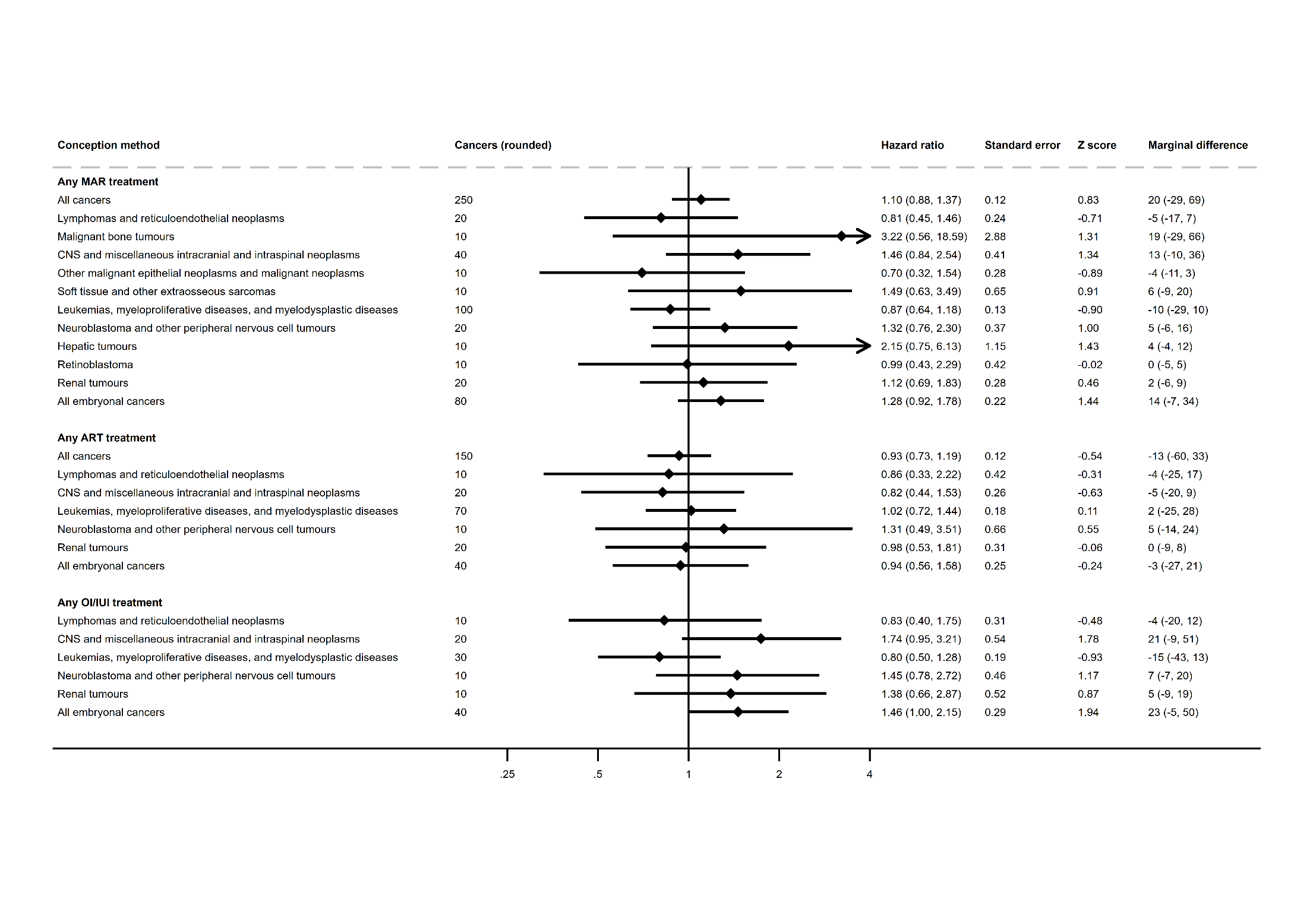

#### Supplementary Figure 8: Standardised incidence ratios (SIRs) splitting assisted reproductive treatments (ART) by in-vitro fertilisation (IVF) and intracytoplasmic sperm injection (ICSI); and by fresh versus frozen embryo transfer – doubly-robust control for maternal age.

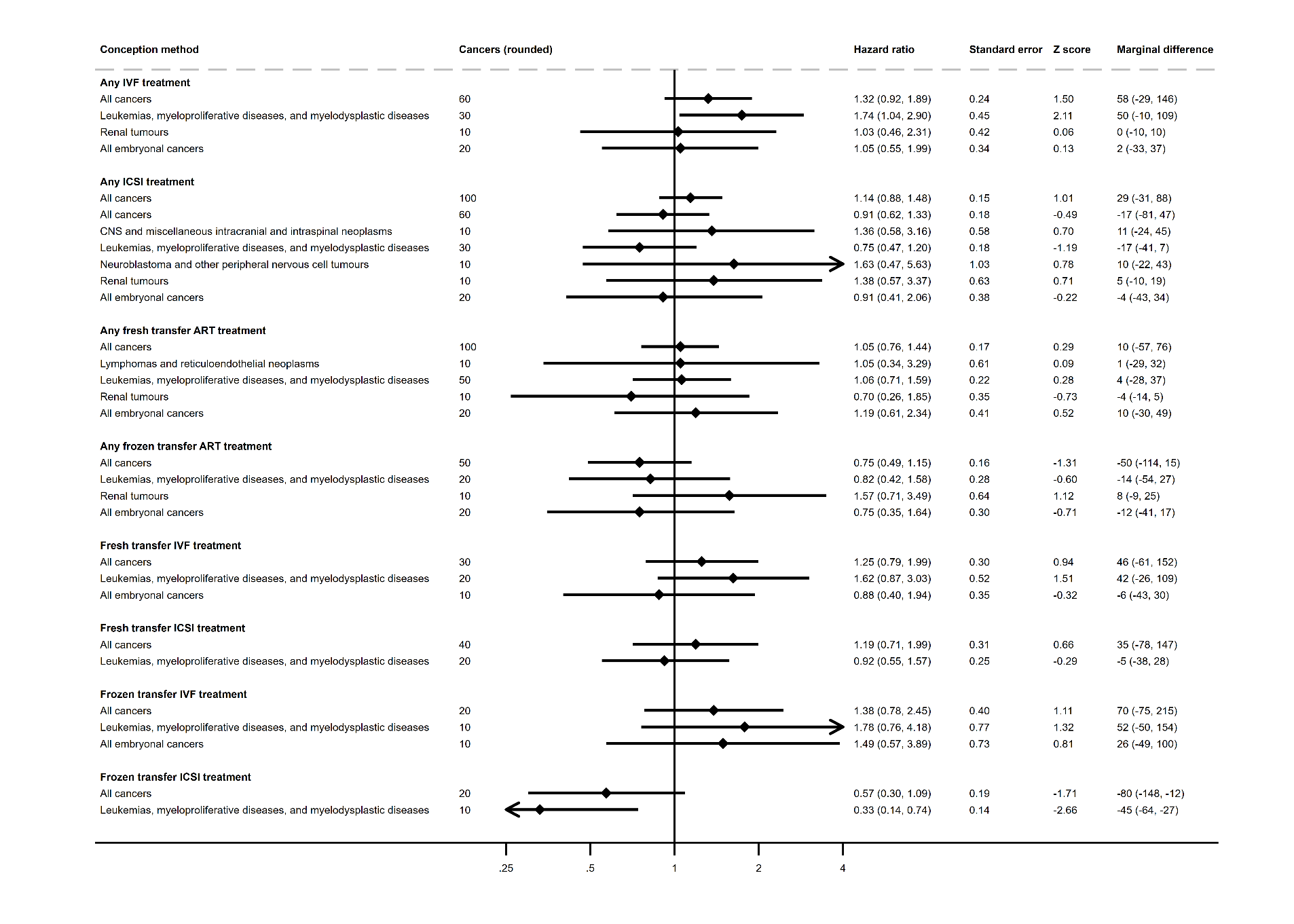
